# Sensorimotor recovery and neuropathic pain reduction after remotely delivered cognitive multisensory rehabilitation or remotely delivered exercise in adults with spinal cord injury: a pilot clinical trial

**DOI:** 10.64898/2026.06.02.26354574

**Authors:** Ann Van de Winckel, Amanda A. Herrmann, Sydney T. Carpentier, Sara Bottale, Ricky L. Lopez, Andrew D. Rapacz, Samantha J. Larson, Wei Deng, Lin Zhang, Timothy J. Hendrickson, Bryon A. Mueller, Ruhollah Nourian, Leslie R. Morse, Kelvin O. Lim

## Abstract

**Introduction:** Reduced or lost sensation and movement after a spinal cord injury (SCI) impairs the brain’s ability to accurately localize paralyzed body parts, causing deficits in its internal body map, or “mental body representations” (MBR). These deficits hinder functional recovery and contribute to neuropathic pain. Medications for neuropathic pain are often ineffective and carry side-effects. Our pilot trials found that *in-person* Cognitive Multisensory Rehabilitation (CMR), a physical therapy restoring MBR, led to prolonged pain reduction, improved sensorimotor function, and enhanced brain function, to greater extent than adaptive fitness.

To explore more accessible interventions for those in rural areas or with transportation challenges, we examined whether 12 weeks of *remotely delivered* CMR or exercise would (1) improve function and reduce pain; (2) increase brain activity and connectivity related to sensorimotor function and MBR in adults with SCI.

**Methods:** Of 19 adults with SCI who consented, 15 (51±15 years old, 8±10 years post-SCI) were randomized to 12 weeks of remotely delivered CMR or exercise (45min, 3x/week). Eight reported neuropathic pain ≥3/10. The Numeric Pain Rating Scale (NPRS), ASIA Impairment Scale (AIS), and Neuromuscular Recovery Scale (NRS) assessed pain and sensorimotor function at baseline, post-intervention, and 6-month follow-up. Functional MRI included resting-state and four tasks: imagining feeling the left leg, imagining moving the left leg, whole-body movement imagery, and a sensation task.

**Results:** After CMR (n=8), participants improved on AIS (large effect sizes: touch: d=1.30; pinprick: d=1.21; lower limb motor function: d=1.83). Exercise (n=7) produced smaller improvements (touch: d=0.35; pinprick: d=0.36; lower limb motor function: d=0.80). CMR showed greater NRS effect sizes (core: d=1.48; upper limb: d=0.69; lower limb: d=1.25) than exercise (core: d=0.31; upper limb: d=0.74; lower limb: d=0.83). Benefits persisted at follow-up for both AIS and NRS, especially in the CMR group. Highest neuropathic pain intensity decreased in both groups post-intervention (CMR: d=-0.61; exercise: d=-0.73) and at 6-month follow-up (CMR: d=-0.55; exercise: d=-0.55). Unlike previous studies, group effects for CMR were not found due to high heterogeneity. Increased task-based activation, including in the lateral occipital cortex involved in visual body perception and spatial awareness, was seen for the exercise group (n=5).

**Discussion:** These preliminary results support the potential of remotely delivered CMR and exercise to improve function and reduce neuropathic pain in adults with SCI, highlighting the need for larger trials.

Clinicaltrial.gov: NCT05870189

## 1 Introduction

Adults with spinal cord injury (SCI) often have partial or complete sensory and motor loss, but in addition about 69% of them experience excruciating neuropathic pain.(1–4) Exercise (with a physical therapist or exercise physiologist) is a common approach to help achieve functional motor recovery. The primary treatment for neuropathic pain is medication, but those are not always efficient and carry risk for significant side effects. A recent systematic review pointed out that the current approaches in research (neuromodulation such as rTMS and tDCS, TENS, virtual reality, cognitive behavioral therapy, mindfulness) did not produce a significant reduction in neuropathic pain. Small pilot studies (n=9-24) in physical activity, visual imagery, hypnosis, osteopathic treatment and acupuncture produced short-term significant effects.(5,6) Therefore, there is a need for non-pharmacological interventions that address both functional recovery and pain to improve quality of life for adults with SCI.

Our previous brain imaging studies and clinical trials in both SCI and stroke have demonstrated that sensory loss contributes to a deficit in mental body representations (MBR), defined as the brain’s internal body map.(7–10) This work confirms that psychological and neuroimaging literature on body schema and body image provides a critical framework for understanding MBR in the context of sensorimotor integration.(3,4,7,8,11–25) Specifically, this collective research demonstrates the distinct roles of these regions within the sensorimotor network: the parietal operculum and insula mediate whole-body awareness and chronic pain perception, whereas the posterior parietal cortex creates the moment-to-moment visuospatial body maps necessary to guide movement.(3,4,11–21) Therefore, awareness of the body and its position in space is inextricably linked to the sensorimotor network, making it an essential component to address during functional recovery and neurorehabilitation.

We have previously demonstrated that SCI leads to weaker sensorimotor network connectivity and maladaptive neuroplasticity, compared to non-injured adults.(8) Without accurate MBR, the brain cannot locate the (paralyzed) limbs, thus, severely limiting motor recovery (e.g., walking).

Cognitive Multisensory Rehabilitation (CMR) is a physical therapy approach that helps adults with neurological disorders restore MBR. We demonstrated in two pilot clinical trial studies (i.e., a delayed-intervention clinical trial and randomized controlled trial comparing CMR to adaptive fitness) increased brain activity during task-based fMRI and increased connectivity, both in areas relevant for MBR and sensorimotor function.(9,22) In the delayed intervention study, the highest neuropathic pain intensity was reduced by 5±3 points, exceeding the minimal clinically important difference (i.e., >1.80 points on the NPRS) for adults with SCI.(26) Sensorimotor improvements were seen for the ISNCSCI ASIA impairment scale (AIS exam), namely touch improved by 9±5 points, pinprick sensation by 8±5 points, and lower limb muscle strength by 4±3 points.

When comparing CMR to adaptive fitness, AIS improved by 7.5±5 points for touch, 8.5±5 points for pinprick and 3±3 points for lower limb strength after CMR (large effect sizes) compared to 2±4 points for touch, 2±4 for pinprick and 1±1.5 points for lower limb strength (small/moderate effect sizes) after adaptive fitness. NRS was increased after CMR by 4±2 points for core strength, 6±9 points for arm function, and 2±2 points for leg function (moderate to large effect sizes) compared to 2±2 points, 1±2.5 points, and 0±0 points, respectively, for the same subscales after adaptive fitness (no to moderate effect sizes). Only 10 out of 16 participants had neuropathic pain and considering the whole group, pain intensity reduced with moderate effect size after CMR and slightly increased after adaptive fitness. Benefits persisted at the 3-month follow-up. CMR increased brain connectivity and activation during the leg imagery task. Increased activation during whole-body imagery was greater after CMR than fitness.

In sum, our two previous pilot clinical trials showed that Cognitive Multisensory Rehabilitation (CMR) produced prolonged reductions in neuropathic pain, improved sensorimotor function, and enhanced brain function, to greater extent than adaptive fitness.

Yet, so far these studies have investigated in-person therapies. Adults with SCI commonly experience transportation issues, a problem that becomes exponentially larger for people living in rural areas. A multisite randomized controlled trial (n=114) comparing 6-month outcomes of telerehabilitation vs standard care showed improvements in function on the functional independence measure (FIM), notably in transfers, in only 1 of the 3 sites (Italy). Another study in Spain in 42 adults with SCI reported no significant difference in FIM outcomes for telerehabilitation vs in person standard care.(27) In general, since COVID-19, telerehabilitation use for adults with SCI in standard care has increased.(28) While telerehabilitation seems to be acceptable and feasible, and early evidence for efficacy for pain and function is found (especially when receiving training from an SCI rehabilitation clinician), more research is needed.(29)

Therefore, the aims of this study were to (1) investigate whether 12 weeks of remotely delivered CMR or remotely delivered exercise would (1) improve sensorimotor function and reduced pain; (2) result in greater brain activity and connectivity related to sensorimotor function and MBR in adults with SCI.

## 2 Materials and methods

### 2.1 Study design

Participants were recruited for this pilot two-arm randomized controlled trial between 6/7/2023 and 1/2/2024. Participants were randomized using a computer-generated allocation sequence for group allocation. To ensure allocation concealment, the sequence was generated and maintained by an independent statistician not involved in participant recruitment or assessment. Raters, the biostatistician, brain technicians and brain imaging analysts were blinded to group allocation. The study followed the Declaration of Helsinki’s principles 2013.(30) The University of Minnesota (UMN)’s Institutional Review Board approved the study, STUDY00018287, on 3/27/2023.

### 2.2 Recruitment

Participants were recruited through clinics and rehabilitation centers in the Twin Cities of the Minnesota Regional Spinal Cord Injury Model System, and clinicaltrials.gov (NCT05870189).

### 2.3 Participants

Inclusion criteria encompassed: Diagnosis of SCI ≥3 months, medically stable, English speaking. Exclusion criteria were ventilator dependency, uncontrolled seizure disorder, severe pressure sores hindering them from sitting for 45 min, cognitive and/or communicative disability (e.g., due to brain injury) precluding learning or following directions, other major medical complications, or pregnancy (current or planned). Participants with MRI contra-indications (stabilizing hardware is typically MRI safe) were enrolled in the trial; however, they did not undergo an MRI.

A broader range of adults with SCI were recruited to target the underlying mechanism of sensorimotor functional recovery, rather than including only those with pain ≥3/10. Nevertheless, given that almost 70% of adults with SCI have neuropathic pain, we prospectively tracked neuropathic pain as an important secondary outcome.

### 2.4 Study procedures

Neuropathic pain was evaluated using the Numeric Pain Rating Scale (NPRS) and the Douleur Neuropathique 4 (DN4),(31) which assesses neuropathic pain by neuropathic pain characteristics. Sensorimotor function was evaluated with the ASIA Impairment Scale (AIS), also called, International Standards for Neurological Classification of Spinal Cord Injury (ISNCSCI), and functional recovery with the Neuromuscular Recovery Scale (NRS). All clinical assessments and behavioral questionnaires were conducted at baseline, post-intervention, and six-month follow-up. Functional magnetic resonance imaging (fMRI) included resting-state and 4 tasks: 1– feeling the left leg; 2– imagining the feeling of moving the left leg, 3– whole-body movement imagery, and 4– a sensation task. MRI testing was performed at baseline and after post-intervention.

AIS and NRS exams were conducted in the wheelchair accessible Brain Body Mind Lab of Dr. Van de Winckel at the UMN. MRI scanning took place at the UMN’s Center for Magnetic Resonance Research (CMRR), with participants screened via a CMRR-prescreening questionnaire. Behavioral questionnaires were administered by trained study staff via secure UMN Zoom. Participant responses were recorded using UMN Research Electronic Data Capture (REDCap), a secure web platform for building and managing online databases and surveys.

### 2.5 Aims

Our first aim was to determine whether 12 weeks of remotely delivered CMR or exercise improved sensory and motor function in adults with SCI, with CMR hypothesized to show greater improvement on performance-based clinical tests of sensorimotor function. Secondary outcomes included neuropathic pain intensity, spasm, mood, self-reported function, body awareness, self-efficacy, sleep, and SCI-related quality of life (namely, positive affect, resilience, self-esteem, ability to participate, pain behavior, and independence), all hypothesized to improve after CMR.

Our second aim was to determine whether 12 weeks of remotely delivered CMR or exercise increased brain activity and connectivity in MBR– and sensorimotor brain areas and networks in adults with SCI, assessed by resting-state and 4 task-based fMRI. We hypothesized that CMR would increase parietal operculum, insula, and posterior cingulate cortex connectivity with other sensorimotor, body awareness, and visuospatial-related brain areas, concurrent with clinical sensorimotor improvements.

### 2.6 Interventions

Both interventions were remotely delivered and provided 1:1, for 12 weeks, 3x/week via UMN Secure Zoom or Microsoft Teams. The sessions were scheduled for 60 min, structured to achieve 45 min of active treatment or exercise. We allowed participants to reschedule missed sessions in the same or following week, e.g. if sessions were missed due to illness or other unforeseen circumstances. All sessions were video-recorded. Logs were created to monitor content, fidelity, and attendance.

Both interventions were classified as minimal risk: remotely delivered CMR due to low-intensity movements and kinesthetic imagery; and remotely delivered exercise as it is part of standard care. It is a standard, community-accessible exercise program, available to adults with SCI following clinical neurorehabilitation.

#### 2.6.1 Remotely delivered Cognitive Multisensory Rehabilitation

The intervention started with a single, brief in-person session during which a certified CMR therapist instructed the participant’s caregiver on how to properly assist with gently moving the participant’s limbs to a specific position. Over the subsequent 12-week intervention, the CMR therapist remotely guided both the caregiver and the participant during live sessions conducted via the UMN Secure Zoom platform. The treating clinician held a formal 3-year post-professional CMR certification awarded by the founding CMR center in Italy (Centro Studi di Riabilitazione Neurocognitiva).

Cognitive Multisensory Rehabilitation (CMR) is a rehabilitation approach, founded in Italy, and specifically focused on restoring MBR to improve daily actions and how the body is interacting with the environment. Based on evidence-based research of neurorehabilitation and neuroscience, the CMR-certified therapist guides the participant to identify body signals, sensations, and use kinesthetic imagery (i.e., imagining the feeling in the body rather than visualizing the body) to solve multisensory discrimination exercises.

The multisensory discrimination exercises are used as a tool to improve MBR, so the questions asked to solve the exercises all relate to the body (e.g., recognizing limb positions in space, or positions of one limb relative to another). Participants also relearn to recognize and integrate correct touch and pressure sensation, because some of the discrimination exercises use different textures or sponges of varying resistance.

Importantly, the key to this MBR restoration is the guidance that the therapist provides, for example, through left-right comparisons, contrasting past memories of actions involving the body with present actions, body scans, comparison of their body position or movements with movements demonstrated by the therapist, or a drawing representing body parts (e.g., the pelvis) to facilitate them to access the representation of that body part. The therapist uses the drawing as a tool to help the participant build awareness of their body in a way they are not doing spontaneously; to let the participant discover their own body weight distribution and then change it into a better one.

The CMR delivery is standardized through a specific reasoning-based therapeutic protocol that the therapist will go through, but the choice of the specific exercise and body part that will be addressed will be personalized to the specific problem of each patient. The therapeutic protocol encompasses understanding where the neuropathic pain is or which movements and actions are particularly difficult (both by observation of actions of the participants and by the participant sharing their experience). The therapist will use this information to create a hypothesis on which MBR would be impaired and will test out this hypothesis through an evaluative exercise. If the participant indeed has difficulty with this exercise, then this exercise will be implemented in the therapy until that aspect of MBR is restored. That will be evaluated after each therapy session by testing that aspect in the action intended to be improved.

Options of body parts that are addressed during CMR related to MBR are 1-dimensions of arms, legs, hands, feet, trunk, and pelvis; 2-the role of the pelvis and feet as the base of support and the sensation of the pelvis as a central body reference, which are important for transfers, balance, and walking; 3-the relationship between body parts, including left and right sides of the body, as well as upper and lower parts of the body; this can be for example, the relationship between the pelvis and the feet.

CMR helps reinvolve the paralyzed body parts in MBR; and multisensory signals become consistently perceived and integrated. Sensory feedback is essential for movement, and thus, sensory gains create the conditions for functional motor recovery. Once the brain accurately maps the body, neuropathic pain and spasms are also reduced or resolved. Concrete examples of CMR exercises in two participants are provided in **Supplementary Data Sheet 1**. Further details can be found in the protocol paper and our prior publication of CMR in adults with SCI-related neuropathic pain.(9,22)

#### 2.6.2. Remotely delivered exercise

Prior to beginning the exercise program, a physiatrist evaluated participants post-baseline at the Brain Body Mind Lab to confirm safety and provide written medical clearance. Remotely delivered exercise sessions were conducted via the Microsoft Teams platform by a team of two certified exercise physiologists at the HealthPartners Rehabilitation, certified through the American College of Sports Medicine (ACSM). Caregiver or partner assistance was permitted to facilitate transfers, equipment setup, or camera adjustments, ensuring the exercise specialists could maintain optimal, real-time remote monitoring throughout each session.

Intervention protocols were designed in accordance with the American College of Sports Medicine guidelines, which recommend that adults with SCI participate in moderate-to-vigorous aerobic exercise 2–3 times per week for ≥20 minutes, resistance training targeting major muscle groups at moderate-to-vigorous intensity, and regular flexibility training.(32) Accordingly, each session consisted of **20 min aerobic exercise, 20 min resistance training, and 5 min of flexibility, core stabilization, and seated balance training**.

Exercise selection, progression, and modification were individualized based on participant functional capacity, neurological presentation, available equipment, individual goals, and observed performance. Exercise intensity was prescribed and monitored using the Borg Rating of Perceived Exertion (RPE) scale, targeting moderate-to-vigorous intensity (RPE 12–16).(33,34) While the exercise physiologists used material that was available in the participant’s homes, our team provided elastic resistance bands (TheraBand®) in three different resistance strengths to all participants in the exercise group to ensure standardized resistance training capabilities. Concrete examples of exercises for two participants are provided in **Supplementary Data Sheet 1**.

Finally, to promote consistency and support intervention fidelity, exercise physiologists recorded subjective responses and objective training parameters for each session using a standardized exercise session log. Intervention delivery and documentation procedures were standardized across participants.

### 2.7 Primary clinical outcomes

The AIS and NRS were performed by a trained clinician or therapist. The AIS (35,36) exam is commonly used in clinics to evaluate the extent and severity of SCI. The subscales of the AIS are light touch (with a cotton ball), pinprick (distinguishing sharp versus dull with a safety pin); and muscle strength testing. Both sensation test scores range from 0 to 54 per side (anus test excluded), and muscle strength for upper and lower limbs have a scoring range of 0 to 25 per limb per side. Higher scores indicate better function or better sensation. The AIS was assessed at baseline, post-intervention, and at the 6-month follow-up.

The NRS (36,37) is used to examine 11 functional tasks, encompassing trunk balance and core strength, arm/hand function, and lower extremity tasks (e.g., sit-to-stand, weight-bearing, taking steps, and stepping over an obstacle). Non-compensatory functional recovery is scored. Higher scores indicate better performance.

### 2.8 Secondary clinical outcomes

The Numeric Pain Rating Scale (NPRS)(38) was used to assess the highest, average (i.e., most of the time), and lowest neuropathic pain intensity in the prior week. The scores range from 0 (no pain) to 10 (worst pain ever). Neuropathic pain was assessed using the Douleur Neuropathique 4 (DN4).(31) This is a diagnostic tool used to identify and assess neuropathic pain by asking about different characteristics of neuropathic pain.(31)

The Patient-Specific Functional Scale (PSFS) encompasses three self-identified functional activities that were important to them. The activities had a measurable goal, scored between 0– unable to do to 10– able to do the activity.(39) The Spinal Cord Injury Functional Index/Assistive Technology (SCI-FI/AT) has 32 self-report items, rated by self-reported difficulty level to perform activities with or without assistive devices, (i.e., 0-“unable to do” to 4-“no difficulty”). The subscales cover basic mobility, self-care, fine motor function, and ambulation.(40,41)

The Penn Spasm Frequency Scale(42) was used to monitor self-reported frequency and severity of muscle spasms. The Pittsburgh Sleep Quality Index (PSQI) contains 7 components. Higher scores indicate worse sleep performance, with a cut-off score of >5 indicating poor sleep quality, and scores ranging from 0 to 21.(43) The Autonomic Standards Assessment form (ANS) is a self-reported questionnaire on cardiovascular health, bladder, bowel and sexual function.(44)

The Spielberger State Trait Anxiety Inventory assesses general feelings (TAI) and present feelings of anxiety (SAI), with scores ranging from 20 to 80, higher scores indicating more anxiety.(45) The Patient Health Questionnaire (PHQ-9) comprises 9 items related to symptoms of depression, with a total scores ranging from 0-27; and higher scores indicating greater symptoms of depression.(46)

The Tampa Scale for Kinesiophobia (TSK) consists of 17 items (total score ranging from 17 to 68) with lower scores indicating greater fear of re-injury.(47) The Moorong Self-Efficacy Scale (MSES) addresses self-reported self-efficacy related to everyday activities for adults with SCI, with a total score range from 16 to 112. Higher scores indicate greater self-efficacy.(48,49) The Kinesthetic and Visual Imagery Questionnaire (KVIQ) assesses the visual imagery clarity from a third person perspective, and kinesthetic imagery intensity of sensations from a first-person perspective. Higher scores reflect better imagery.(50)

The Functionality Appreciation Scale (FAS, 7 items) identifies how appreciative a person is for their body functionality.(51,52) The Postural Awareness Scale (PAS, 12 items) assesses self-reported awareness of body posture.(53) The Revised Body Awareness Rating Questionnaire (BARQ-R, 12 items) evaluates body states, e.g., how tension in the body affects one’s body awareness and function in daily life. Lower scores indicate better body awareness.(54,55)

The Multidimensional Assessment Interoceptive Awareness – version 2 (MAIA-2, 37 items) measures 8 domains related to interoception. The domains encompass: *noticing* (awareness of body sensations); *not-distracting* (tendency to not ignore/distract oneself from sensations of pain or discomfort); *not-worrying* (tendency not to worry or experience emotional distress with sensations of pain or discomfort); *attention regulation* (ability to sustain attention to body sensations); *emotional awareness* (awareness of the connection between body sensations and emotional states); *self-regulation* (ability to regulate distress by attention to body sensations); *body listening* (active listening to the body for insight), and *trusting* (experience of one’s body as safe and trustworthy).(56)

The Spinal Cord Injury Quality of Life measurement system (SCI-QOL) evaluates 4 broad domains (22 subscales) of physical/medical health, emotional health, social participation, and physical functioning. We used 6 short-form subscales: positive affect (10 items), resilience (8 items), self-esteem (8 items), ability to participate (10 items), independence (8 items), pain behavior (7 items).(57)

The Craig Handicap Assessment and Reporting Technique-Short Form (CHART-SF) assesses the degree by which a person with SCI remains with an impairment or disability. It encompasses the type and level of assistance needed physically and cognitively; mobility (i.e., level of physical activity and transportation needs); occupation (how time is spent); social interactions; and financial resources (economic self-sufficiency). Each of six domains is scored from 0-100, with total scores ranging from 0-600. Higher scores indicate less impairment.(58)

Participants attended weekly (during the intervention) and monthly (during the follow-up) phone check-ins with a research assistant who recorded any adverse events (also those unrelated to the study), content and perceived effects of the intervention, use of over-the-counter or prescribed medications for spasms or neuropathic pain, if present, recent illnesses, healthcare utilization and/or recent hospitalizations, and information related to community integration. We allowed concomitant medication prescribed or recommended by their clinical care team throughout the trial, as well as other treatments/therapies related to their standard of care such as stretching. However, participants did not receive additional balance, gait, or strengthening exercise outside of the study interventions, or any additional pain management treatments aside from prescribed or over the counter medication.

### 2.9 Brain imaging outcomes

Participants were scanned for 2 hours on a Siemens 3-T Prisma scanner at CMRR. Full acquisition details are reported in a previous protocol paper.(9)

In short, structural MRI acquisition included T1-weighted magnetization-prepared rapid acquisition with gradient echo (MPRAGE) [repetition time (TR)=2.5s; echo time (TE)=4.5ms; 0.8mm isotropic voxels], and T2-weighted sampling perfection with application-optimized contrasts using different flip angle evolution (SPACE) [TR=3.2s; TE=565ms; 0.8mm isotropic voxels].

The resting-state scan lasted for 12 min 10 sec. Participants were asked to maintain eye fixation with a restful mind. Both resting-state and task-based fMRI scans were acquired with T2*-weighted multiband echo planar acquisition tilted 30 degrees relative to the anterior commissure–posterior commissure (AC-PC) plane according to auto-align software, enabling whole-head blood-oxygen-level-dependent (BOLD)–contrast with optimal temporal and spatial resolution and to reduce signal dropout [TR = 0.8 s; TE = 37ms; flip angle = 55 degrees; 72 slices; multiband factor 8; 2 mm isotropic voxels].

Of particular interest were brain areas and networks involved in sensorimotor and MBR function in adults with SCI. Therefore, seeds in the parietal operculum, insula, and posterior cingulate cortex were selected based on our CMR work in stroke and SCI, as well as other research demonstrating their importance in MBR.(7–10)

**Task 1: Sensory imagery of the left leg (8 min 15 sec)**. This task requires participants to direct a gentle attention toward any emerging sensations within the affected left leg. This process of mindful somatic scanning is alternated with rest. This task stems directly from our previous research in adults with chronic low back pain, in which individual changes were noted of increased parietal operculum and the posterior parietal cortex activation.(59) Increased insular activity was also seen in another study during mindful body scanning.(60)

**Task 2: Kinesthetic imagery of the left leg (7 min 58 sec).** This task protocol alternated blocks of rest with periods of active imagery, during which participants maintained a gentle, non-judgmental awareness of the feeling in the body of imagining moving the left leg (without actually moving). In prior research, mindfulness meditation strategies have demonstrated increased insula and parietal operculum activation.(61)

**Task 3: Whole-body movement imagery (19 min 51 sec).** Participants first watch a demonstration video of a Qigong (whole-body) movement, and then, without executing any actual physical motion, perform a kinesthetic imagery of this movement.(62) The video showed a Qigong master standing with legs at shoulder width, breathing in, bending the knees and opening the arms gently, followed by breathing out, straightening the knees and bringing the arms closer. Neuroimaging literature confirming that motor imagery and actual motor execution share highly overlapping spatial networks in the brain.(63–66) Furthermore, our prior research verified that this exact full-body sequence drives activation within the posterior parietal cortex and parietal operculum in adults with chronic low back pain.(59)

**Task 4: Sensory stimulation of the big toes and thumbs (18 min 32 sec).** The pads of the participant’s big toes and thumbs were gently stroked with a towel. The participant was not told beforehand which specific extremity would be targeted. Once the scan concludes, participants are asked to verify if they felt the strokes and, if so, to identify their exact locations. This task was chosen because the same task in our previous study with adults with SCI-related neuropathic pain demonstrated greater activation in the bilateral postcentral gyrus (mediating somatosensory processing); the angular gyrus, supramarginal gyrus, and superior parietal lobe (components of the posterior parietal cortex responsible for visuospatial body maps); the left insula (involved in bodily awareness and pain processing); and the frontal operculum.(22)

### 2.10 Statistical analysis

#### 2.10.1 Clinical outcome analysis

<u>Sample size calculation:</u> For Aim 1, based on our prior data of CMR in adults with SCI, we estimate an average 12-week sensorimotor function improvement of 7.50±4.89 points for remotely delivered CMR. A sample size of 7 participants will have about 81% power to detect mean pre-post differences of effect size d=1.5 for the remote CMR or remote exercise in sensorimotor function after 12 weeks at an overall Type 1 error rate of α=0.05 with Bonferroni correction.

For Aim 2 (n=11/group), we will have about 84% power to detect mean pre-post changes of connectivity strength or brain activation of an effect size d=1.25 for remote CMR or remote exercises with an overall Type 1 error rate of α=0.05 with Bonferroni correction. We will have 80% power to detect a correlation of 0.74 between sensorimotor function and brain function changes post-intervention for each group.

The present data are essential to provide preliminary evidence of remote intervention’s effectiveness to support a future larger randomized controlled trial.

We summarized all quantitative variables with descriptive statistics at each time point per group. We used R for all statistical analyses. Given the small sample size, we reported Cohen’s d effect sizes rather than p-values for within-group pre-post changes and pre-6-month follow-up changes in sensorimotor function measures and other clinical outcomes.(67)

A UMN Clinical and Translational Science Institute data integrity monitor and quality assurance reviewer monitored the study every 6 months.

#### 2.10.2 Brain imaging analysis

Initial data preprocessing for all neuroimaging runs was conducted using the Human Connectome Project (HCP) pipeline. Subsequent fMRI processing was carried out via the CONN functional connectivity toolbox.(68) Data underwent realignment, scrubbing, and artifact detection, followed by CompCor denoising. We applied established family-wise error correction protocols based on Statistical Parametric Mapping, to control for multiple comparisons. The preprocessing sequence was uniformly applied to both the task-based and resting-state fMRI datasets.

Following preprocessing, individualized voxel-wise brain activation for the task-based fMRI data was modeled using a standard general linear model. For the resting-state data, our primary analytic approach consisted of a voxel-wise, whole-brain seed-based functional connectivity analysis derived from Pearson correlation coefficients. This connectivity analysis specifically targeted the insula, parietal operculum, and posterior cingulate cortex as a priori regions of interest, and was conducted alongside exploratory multivariate pattern analyses.

To evaluate within-group shifts from baseline to post-intervention, resting-state connectivity values (converted via Fisher’s Z transformation) and task-evoked activation maps were analyzed using appropriate paired t-tests or Wilcoxon signed-rank tests.

We tested the effect of CMR on brain imaging outcomes using mixed-effects models implemented in FSL. Cluster-based correction for multiple comparisons was performed using a cluster-forming threshold of Z > 1.96 and a cluster-level significance threshold of p < 0.05 to control the family-wise Type I error rate.

## 2 Results

**Fig. 1** displays the CONSORT flow diagram. Of the 19 participants screened for eligibility and who consented, 4 participants with SCI withdrew: 3 because they realized they could not commit to the intervention because of work engagements; one stopped communicating after signing consent. We collected MRI scans for 11 participants at baseline. Reasons for not having an MRI were either epidural stimulators (n= 2), too much hardware in the body (n=1) or a bullet fragment in the body (n=1).

**Fig. 1.**
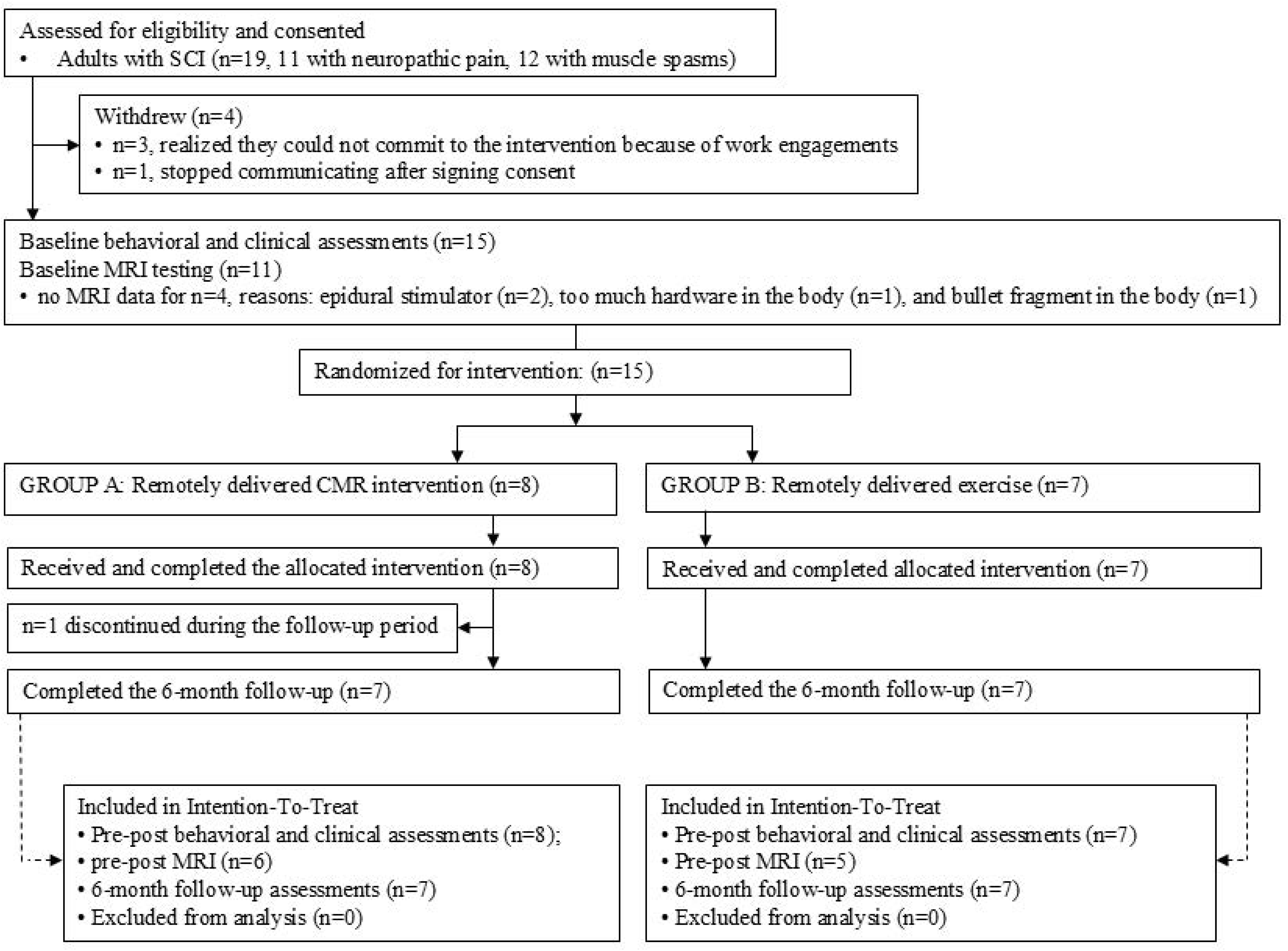
CONSORT flow diagram.

Fifteen adults were randomized to the interventions. The group had an average age of 51 ± 15 years (range of 25-72 years), 9 women, and 6 men; All White non-Hispanic adults, 2 Veterans, 47% living in rural areas, 53 % married, all with at least some college education. Of the 8 participants who had neuropathic pain at baseline, their highest neuropathic pain level was 6 ± 3 on the NPRS. They were 8 ± 10 years after SCI (range of 0.5-38.5 years). Six adults had tetraplegia, and 9 had paraplegia, 3 had a complete SCI. There were no significant baseline differences in AIS or NRS outcomes between the groups (*p*-values: 0.22-0.73). The demographic and clinical information of the 15 participants are detailed in **Table 1**.

All 8 participants completed the *remotely delivered CMR intervention.* One person withdrew during the follow-up phase, and, thus, 7 completed the study. Seven participants started the *remotely delivered exercise intervention*. All 7 participants completed the study. All participants averaged 135 min/week (3 sessions of 45min/week) of intervention throughout the 12 weeks. On occasion, sessions were rescheduled in the following week, due to illness. The final analysis included 15 participants in total for the pre-post intervention analysis (n=8 in the CMR group; 7 in the exercise group) and 14 participants for the pre-6-month follow-up analysis (n=7 in the CMR group; 7 in the exercise group).

### 3.1 Clinical outcomes

Clinical outcomes are reported in **Table 2** with effect sizes for post-intervention and 6-month follow-up vs baseline.

Sensorimotor improvements were greater after CMR than after exercise. Participants in the CMR group obtained large effect sizes for AIS (touch: change score 3.38±2.60, CI95=[1.20; 5.55], d=1.30; pinprick: change score 4.69±3.89, CI95=[1.44; 7.94], d=1.21; lower limb motor function: change score 1.88±1.03, CI95=[1.02; 2.73], d=1.83) compared to small-to-large for exercise (touch: change score 1.07±3.03, CI95=[-1.73; 3.88], d=0.35; pinprick: change score 1.00±2.78, CI95=[-1.58; 3.58], d=0.36; lower limb motor function: change score 0.21±0.27, CI95=[-0.03; 0.46], d=0.80, **Fig. 2**). Effect sizes for NRS were also larger after CMR (core: change score 7.75±5.23, CI95=[3.38; 12.12], d=1.48; arm function: change score 3.88±5.64, CI95=[-0.84; 8.59], d=0.69; leg function: change score 5.88±4.70, CI95=[1.94; 9.81], d=1.25) than after exercise (core: change score 0.43±1.40, CI95=[-0.86; 1.72], d=0.31; upper limb: change score 1.57±2.13, CI95=[-0.40; 3.54], d=0.74; lower limb: change score 1.71±2.06, CI95=[-0.19; 3.62], d=0.83, **Fig. 3**).

**Fig. 2.**
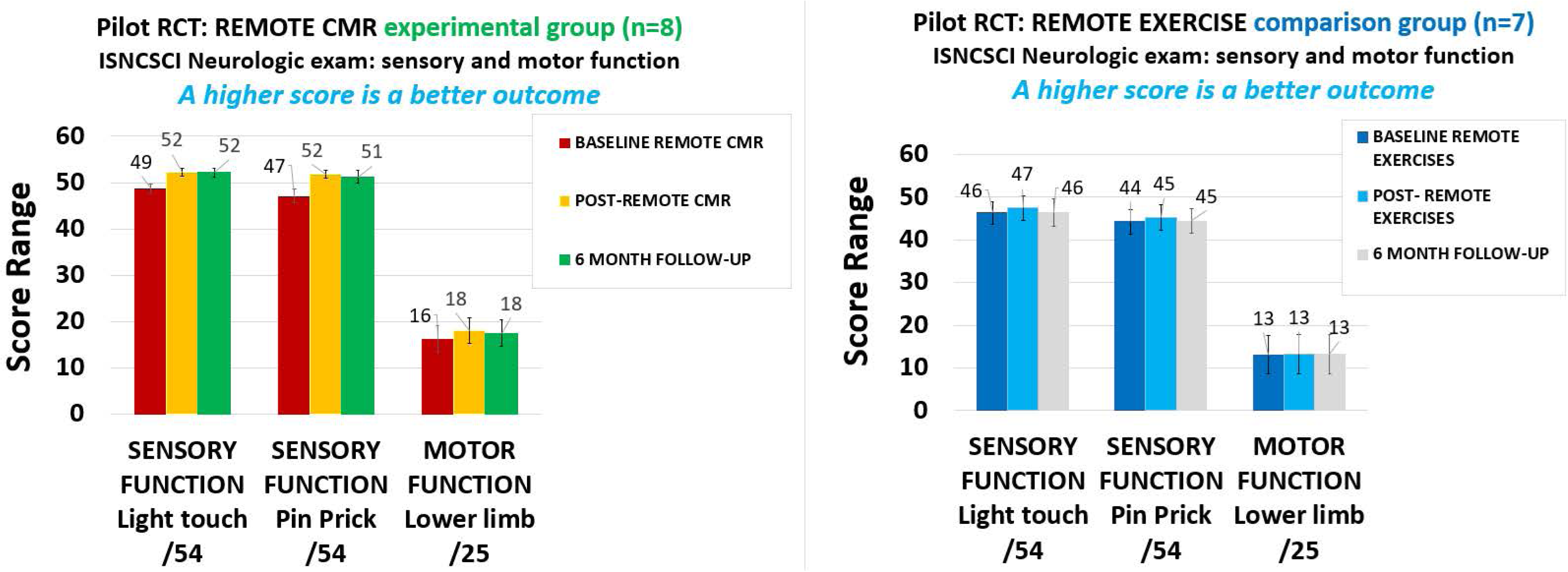
AIS exam. Bar graphs show average left and right scores for touch, pinprick sensation, and lower limb strength at baseline, post-intervention, and 6-month follow-up for the remotely delivered CMR and remotely delivered exercise groups.

**Fig. 3.**
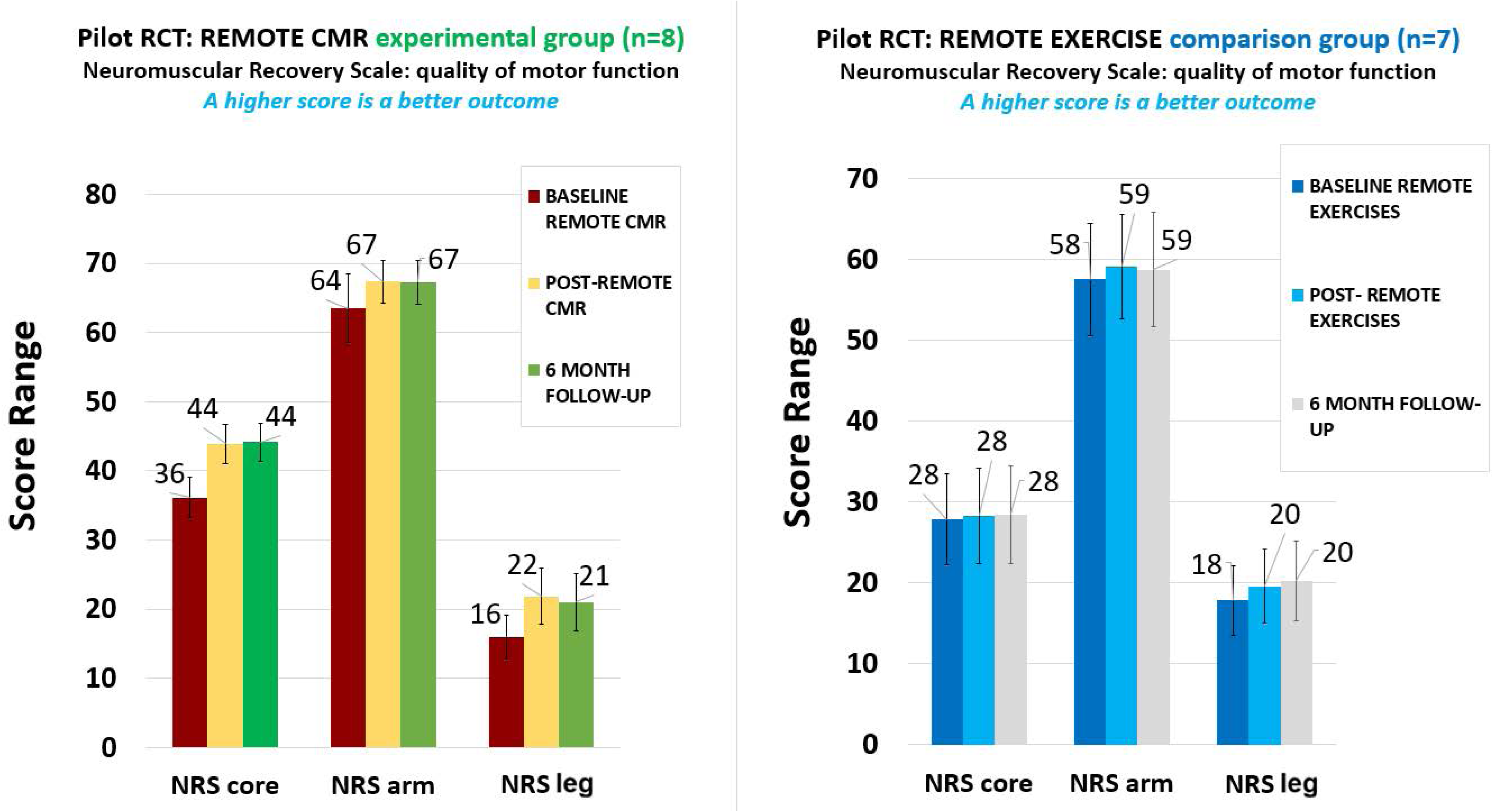
NRS exam. Bar graphs show scores for core strength, arm function, and leg function at baseline, post-intervention, and 6-month follow-up for the remotely delivered CMR and remotely delivered exercise groups.

At 6-month follow-up compared to baseline, CMR maintained large AIS effect sizes (touch: change score 2.93±1.69, CI95=[1.36; 4.50], d=1.73; pinprick: change score 3.93±3.60, CI95=[0.60; 7.26], d=1.09; lower limb motor function: change score 2.29±1.11, CI95=[1.26; 3.32], d=2.05, **Fig. 2**) and moderate-to-large NRS effect sizes (core: change score 7.29±4.23, CI95=[3.37; 11.20], d=1.72; arm function: change score 3.93±6.75, CI95=[-2.31; 10.17], d=0.58; standing: change score 6.86±4.98, CI95=[2.25; 11.46], d=1.38). The exercise group showed no effect-to-moderate AIS effect sizes (touch: change score 0.07±3.92, CI95=[-3.55; 3.70], d=0.02; pinprick: change score 0.29±3.09, CI95=[-2.58; 3.15], d=0.09; lower limb motor function: change score 0.21±0.39, CI95=[-0.15; 0.58], d=0.55) and small-to-large NRS effect sizes (core: change score 0.57±1.81, CI95=[-1.11; 2.25], d=0.32; arm function: change score 1.21±2.60, CI95=[-1.19; 3.62], d=0.47; leg function: change score 2.43±2.76, CI95=[-0.12; 4.98], d=0.88, **Fig. 3**).

Only 8 of 15 participants (i.e., 3 in the CMR group; 5 in the exercise group) had neuropathic pain at baseline (highest pain: 6±3 points on the NPRS), and with large NPRS standard deviations, reflecting substantial inter-individual variability.

When calculating the within-group differences per group for the whole cohort (including those without neuropathic pain), CMR (n=8) showed small-to-moderate post-intervention effect sizes for pain reduction (highest NPRS: change score –1.25±2.05, CI95=[-2.97; 0.47], d=-0.61; average NPRS: change score –0.88±1.81, CI95=[-2.39; 0.64], d=-0.48) and small-to-moderate effects at 6-month follow-up, compared to baseline (highest NPRS: change score –0.86±1.57, CI95=[-2.31; 0.60], d=-0.55; average NPRS: change score –0.29±0.76, CI95=[-0.99; 0.41], d=-0.38).

CMR also reduced the severity of neuropathic pain characteristics (DN4) with a large effect size post-CMR (change score –1.13±1.25, CI95=[-2.17; –0.08], d=-0.90) and a moderate effect size when comparing 6-month follow-up vs baseline (change score –1.00±1.41, CI95=[-2.31; 0.31], d=-0.71).

The exercise group showed small-to-moderate effect sizes for highest and average neuropathic pain on the NPRS from both baseline to post-exercise and baseline to 6-month follow-up: (highest post-exercise: change score –0.57±0.79, CI95=[-1.30; 0.16], d=-0.73; 6-month follow-up: change score – 0.43±0.79, CI95=[-1.16; 0.30], d=-0.55; average post-exercise: change score –0.71±1.25, CI95=[-1.87; 0.45], d=-0.57; 6-month follow-up: change score –0.29±0.76, CI95=[-0.99; 0.41], d=-0.38).

Notably, unlike the CMR group, the remote exercise group did not demonstrate changes in DN4 scores after the exercise intervention and even showed a small increase in neuropathic pain characteristics at 6-month follow-up (change score 0.14±0.38, CI95=[-0.21; 0.49], d=0.38).

#### 3.1.1 Function

Both groups showed large PSFS effect sizes for self-identified functional goal attainment (CMR: change score 4.58±2.59, CI95=[2.42; 6.75], d=1.77; exercise: change score 3.48±2.71, CI95=[0.97; 5.99], d=1.28), which were sustained at follow-up (CMR: change score 5.10±2.58, CI95=[2.71; 7.48], d=1.98; exercise: change score 3.62±2.83, CI95=[1.00; 6.24], d=1.28).

For SCI-FI/AT paraplegia, CMR (n=5) showed large effect sizes at both time points (post-CMR and 6-month follow-up) for basic mobility (change score 3.80±2.49, CI95=[0.71; 6.89], d=1.53; change score 4.40±3.21, CI95=[0.42; 8.39], d=1.37), self-care (change score 3.00±2.74, CI95=[-0.40; 6.40], d=1.10; change score 3.40±2.51, CI95=[0.28; 6.52], d=1.36), fine motor function (change score 1.00±1.23, CI95=[-0.52; 2.52], d=0.82; change score 1.00±1.23, CI95=[-0.52; 2.52], d=0.82), and ambulation (change score 4.00±1.73, CI95=[1.85; 6.15], d=2.31; change score 4.40±2.19, CI95=[1.68; 7.12], d=2.01).

Exercise (n=4) resulted in moderate effect sizes at those two time points: basic mobility (change score 1.00±1.41, CI95=[-1.25; 3.25], d=0.71; change score 1.00±1.41, CI95=[-1.25; 3.25], d=0.71), self-care worsened at follow-up (change score 0.25±0.50, CI95=[-0.55; 1.05], d=0.50; change score – 0.25±0.50, CI95=[-1.05; 0.55], d=-0.50), fine motor function (change score 0.25±0.50, CI95=[-0.55; 1.05], d=0.50; change score 0.25±0.50, CI95=[-0.55; 1.05], d=0.50), and ambulation (change score 0.75±1.50, CI95=[-1.64; 3.14], d=0.50; change score 0.75±1.50, CI95=[-1.64; 3.14], d=0.50).

For the SCI-FI/AT assessment in adults with quadriplegia, baseline data were collected for three participants in each group. At follow-up, all three participants in the exercise group completed the assessment, whereas only two participants in the CMR group completed it. CMR showed large effect sizes post-intervention and at 6-month follow-up for basic mobility (change score 11.33±8.02, CI95=[-8.59; 31.26], d=1.41; change score 9.00±5.66, CI95=[-41.83; 59.83], d=1.59), self-care (change score 6.33±4.73, CI95=[-5.41; 18.07], d=1.34; change score 7.00±4.24, CI95=[-31.12; 45.12], d=1.65), fine motor function (change score 5.00±3.61, CI95=[-3.96; 13.96], d=1.39; change score 6.50±3.54, CI95=[-25.27; 38.27], d=1.84), and ambulation (change score 6.67±3.06, CI95=[-0.92; 14.26], d=2.18; change score 7.00±4.24, CI95=[-31.12; 45.12], d=1.65).

In contrast, no effect to moderate effect sizes were seen after exercise for the two timepoints, with basic mobility even slightly worsening at 6-month follow-up. The values were: Basic mobility (change score 0.00±0.00, d=0.00; change score –0.33±0.58, CI95=[-1.77; 1.10], d=-0.58); self-care (change score 0.33±0.58, CI95=[-1.10; 1.77], d=0.58; change score 0.00±0.00, d=0.00), fine motor function (change score 0.67±1.15, CI95=[-2.20; 3.54], d=0.58; change score 0.00±0.00, d=0.00), and ambulation (change score 0.33±0.58, CI95=[-1.10; 1.77], d=0.58; change score 0.33±0.58, CI95=[-1.10; 1.77], d=0.58).

Mobility, measured with CHART-S, reflected moderate gains following CMR post-intervention (change score 12.13±22.97, CI95=[-7.08; 31.33], d=0.53) and at 6-month follow-up (change score 3.71±5.50, CI95=[-1.37; 8.80], d=0.68), and small gains for exercise at both timepoints (post-intervention: change score 1.43±3.78, CI95=[-2.07; 4.92], d=0.38; 6-month follow-up: change score 1.43±3.78, CI95=[-2.07; 4.92], d=0.38). Participants in the CMR group also experienced moderate improvements in occupation at both timepoints (post-intervention: change score 16.33±31.09, CI95=[-9.67; 42.32], d=0.53; 6-month follow-up: change score 20.27±31.98, CI95=[-9.31; 49.85], d=0.63). Physical independence had some small gains in both groups post-intervention (CMR: change score 2.00±5.66, CI95=[-2.73; 6.73], d=0.35; exercise: change score 2.29±6.05, CI95=[-3.31; 7.88], d=0.38) and at 6-month follow-up (CMR: change score 2.29±6.05, CI95=[-3.31; 7.88], d=0.38; exercise: change score 2.29±6.05, CI95=[-3.31; 7.88], d=0.38). Economic self-sufficiency only had a small improvement at 6-month follow-up in the CMR group (change score 3.23±8.55, CI95=[-4.68; 11.14], d=0.38), with no changes observed post-intervention or in the exercise group.

#### 3.1.2 Body awareness and motor imagery

For KVIQ, 50% of participants in the CMR group already had a maximum score at baseline. Visual and kinesthetic imagery improved from baseline to post-CMR (change score 0.88±1.81, CI95=[-0.64; 2.39], d=0.48) and at 6-month follow-up (change score 1.00±1.92, CI95=[-0.77; 2.77], d=0.52). The exercise group also demonstrated improvements in imagery ability at both post-intervention and 6-month follow-up with identical large effect sizes at both timepoints (change score 2.71±2.56, CI95=[0.34; 5.09], d=1.06).

For functional appreciation (FAS), participants in the CMR group demonstrated large improvements post-intervention (change score 0.50±0.48, CI95=[0.10; 0.90], d=1.04) and at 6-month follow-up (change score 0.47±0.38, CI95=[0.12; 0.82], d=1.25). Conversely, the exercise group showed small improvements post-intervention (change score 0.16±0.75, CI95=[-0.53; 0.86], d=0.22) and moderate improvements at 6-month follow-up (change score 0.36±0.70, CI95=[-0.38; 1.10], d=0.51).

Postural awareness (PAS) improved substantially in the CMR group, demonstrating large effect sizes both post-intervention (change score 8.38±7.48, CI95=[2.12; 14.63], d=1.12) and at 6-month follow-up (change score 7.14±4.88, CI95=[2.63; 11.66], d=1.46). The exercise group also showed enhancements in postural awareness, though with moderate effect sizes across both time points (post-intervention: change score 3.86±7.27, CI95=[-2.86; 10.58], d=0.53; 6-month follow-up: change score 3.14±4.67, CI95=[-1.18; 7.46], d=0.67).

For body awareness (BARQ-R), encompassing body tension, emotional body awareness, and awareness of breathing, with lower scores demonstrating better body awareness, the CMR group showed a small effect size post-intervention (change score –1.50±3.07, CI95=[-4.07; 1.07], d=-0.49), which then returned to baseline at 6-month follow-up (change score 0.14±3.02, CI95=[-2.65; 2.94], d=0.05). In contrast, the exercise group demonstrated large improvements in body awareness post-intervention (change score –1.57±1.13, CI95=[-2.62; –0.52], d=-1.39) and small improvements at 6-month follow-up (change score –1.14±3.44, CI95=[-4.32; 2.04], d=-0.33).

MAIA-2 subscales revealed meaningful enhancements across both cohorts. Noticing abilities improved with a moderate effect size in both the remote CMR group (change score 0.56±0.73, d=0.77) and the remote exercise group (change score 0.68±0.88, d=0.78). Body listening showed a small improvement in the CMR group (d=0.49) and a large improvement in the exercise group (change score 0.62±0.62, CI95=[0.04; 1.19], d=1.00). Similarly, trust improved with a small effect size in the CMR group (d=0.37) and with a large effect size in the exercise group (change score 0.57±0.57, CI95=[0.05; 1.10], d=1.01).

For multidimensional body awareness (MAIA-2), scores revealed varied results across subscales. Post-intervention, both groups demonstrated moderate improvements for the noticing subscale (CMR: change score 0.56±0.73, CI95=[-0.05; 1.17], d=0.77; exercise: change score 0.68±0.88, CI95=[-0.13; 1.49], d=0.78). At 6-month follow-up, the exercise group sustained a large improvement in noticing (change score 0.64±0.66, CI95=[0.03; 1.25], d=0.98), while the CMR group demonstrated a large improvement in not-worrying (change score 0.63±0.59, CI95=[0.08; 1.18], d=1.06). For the body listening subscale, the exercise group demonstrated large improvements at both time points (post-intervention: change score 0.62±0.62, CI95=[0.04; 1.19], d=1.00; 6-month follow-up: change score 0.86±0.42, CI95=[0.47; 1.25], d=2.02). In contrast, the CMR group showed a small improvement post-intervention and a moderate improvement at follow-up for body listening (post-intervention: change score 0.54±1.10, CI95=[-0.38; 1.46], d=0.49; 6-month follow-up: change score 0.52±1.00, CI95=[-0.40; 1.45], d=0.53). Finally, the exercise group showed improvements in trusting that were large post-intervention and moderate at follow-up (post-intervention: change score 0.57±0.57, CI95=[0.05; 1.10], d=1.01; 6-month follow-up: change score 0.62±0.83, CI95=[-0.14; 1.38], d=0.75).

#### 3.1.3 Spasticity, autonomic function, sleep, and psychosocial wellbeing

For spasm frequency, the remote CMR group showed a small reduction post-intervention (change score –0.50±1.07, CI95=[-1.39; 0.39], d=-0.47) and a small reduction at 6-month follow-up (change score –0.14±0.38, CI95=[-0.49; 0.21], d=-0.38). In contrast, the remote exercise group showed no change in spasm frequency at either time point (post-intervention and 6-month follow-up have both change score 0.00±0.00, d=0.00).

For spasm severity, the CMR group demonstrated moderate reductions both post-intervention (change score –0.38±0.74, CI95=[-1.00; 0.25], d=-0.50) and at 6-month follow-up (change score – 0.43±0.79, CI95=[-1.16; 0.30], d=-0.55). The exercise group experienced a small reduction in spasm severity post-intervention (change score –0.14±0.38, CI95=[-0.49; 0.21], d=-0.38), which progressed to a moderate reduction at 6-month follow-up (change score –0.29±0.49, CI95=[-0.74; 0.17], d=-0.59).

For autonomic function (ANS) where higher scores indicate better function, the remote CMR group demonstrated moderate improvements both post-intervention (change score 1.13±1.55, CI95=[-0.17; 2.42], d=0.73) and at 6-month follow-up (change score 1.14±1.77, CI95=[-0.50; 2.78], d=0.65). In contrast, the remote exercise group showed small improvements at both time points (post-intervention: change score at both timepoints compared to baseline were 0.14±0.38, CI95=[-0.21; 0.49], d=0.38).

Both groups experienced better sleep post-intervention. The remote CMR group demonstrated moderate improvements post-intervention (change score –2.63±4.44, CI95=[-6.34; 1.09], d=-0.59) and at 6-month follow-up (change score –2.43±4.12, CI95=[-6.24; 1.38], d=-0.59). The remote exercise group achieved a large improvement post-intervention (change score –1.86±1.57, CI95=[-3.31; –0.40], d=-1.18), but the effect diminished, returning to baseline levels (change score – 0.14±2.04, CI95=[-2.03; 1.74], d=-0.07).

Symptoms of depression (PHQ-9) showed varied results across the time points. Post-intervention, the remote CMR group demonstrated a moderate reduction in symptoms (change score –1.75±2.82, CI95=[-4.10; 0.60], d=-0.62), whereas the exercise group showed no change (change score 0.14±2.34, CI95=[-2.02; 2.31], d=0.06). However, the initial effect for the CMR group did not persist at 6-month follow-up, returning to near baseline levels with no meaningful effect (change score – 0.14±4.45, CI95=[-4.26; 3.97], d=-0.03). Conversely, the remote exercise group demonstrated a delayed small improvement at 6-month follow-up (change score –0.57±2.37, CI95=[-2.76; 1.62], d=-0.24).

For state anxiety (lower scores indicate less anxiety in the moment), the remote CMR group demonstrated a small improvement post-intervention (change score –1.25±4.65, CI95=[-5.14; 2.64], d=-0.27) and a large improvement at 6-month follow-up (change score –4.25±4.20, CI95=[-7.76; – 0.74], d=-1.01). Conversely, the remote exercise group experienced a small worsening in state anxiety post-intervention (change score 0.86±2.73, CI95=[-1.67; 3.39], d=0.31), which returned to baseline levels with no effect at 6-month follow-up (change score 0.00±4.00, CI95=[-3.70; 3.70], d=0.00).

Similarly, trait anxiety (predisposition) decreased in the CMR group, showing small improvements both post-intervention (change score –2.00±4.28, CI95=[-5.58; 1.58], d=-0.47) and at 6-month follow-up (change score –2.57±5.77, CI95=[-7.91; 2.76], d=-0.45). The exercise group showed a small worsening in trait anxiety post-intervention (change score 0.71±3.40, CI95=[-2.43; 3.86], d=0.21), followed by a small improvement at 6-month follow-up (change score –1.14±2.91, CI95=[-3.84; 1.55], d=-0.39).

Kinesiophobia (TSK) showed a small reduction in fear of pain during movement post-CMR (change score –1.13±4.97, CI95=[-5.28; 3.03], d=-0.23), followed by a near baseline score at 6-month follow-up compared to baseline (change score 0.57±4.47, CI95=[-3.56; 4.70], d=0.13). The remote exercise group demonstrated an increase in kinesiophobia, showing a small worsening at both post-intervention (change score 2.43±5.80, CI95=[-2.93; 7.79], d=0.42) and at 6-month follow-up (change score 1.43±3.41, CI95=[-1.72; 4.58], d=0.42).

The MSES demonstrated consistent enhancements in self-efficacy across both intervention arms. The remote CMR group showed a small improvement post-intervention (change score 2.13±6.47, CI95=[-3.28; 7.53], d=0.33), which was maintained at 6-month follow-up (change score 1.43±4.79, CI95=[-3.00; 5.86], d=0.30). Similarly, the remote exercise group exhibited a small improvement post-intervention (change score 1.57±5.47, CI95=[-3.49; 6.63], d=0.29) and at 6-month follow-up (change score 0.86±2.61, CI95=[-1.56; 3.27], d=0.33).

For SCI-QOL subscales, notable changes included a small improvement in positive affect for the remote CMR group at 6-month follow-up (change score 2.03±4.19, CI95=[-1.84; 5.90], d=0.49). Resilience demonstrated small improvements post-CMR (change score 1.95±6.80, CI95=[-3.73; 7.63], d=0.29) and at 6-month follow-up in both groups (CMR: change score 2.39±6.78, CI95=[-3.88; 8.66], d=0.35; exercise: change score 1.15±4.72, CI95=[-3.81; 6.11], d=0.24). Ability to participate also improved in both groups, with small changes post-intervention (CMR: change score 2.43±9.16, CI95=[-5.23; 10.08], d=0.27; exercise: change score 1.63±5.52, CI95=[-3.47; 6.73], d=0.30) and small-to-moderate changes at 6-month follow-up (CMR: change score 4.31±10.22, CI95=[-5.14; 13.76], d=0.42; exercise: change score 1.25±1.87, CI95=[-0.71; 3.21], d=0.67).

Conversely, certain subscales showed varied outcomes for the exercise group. Pain behavior decreased (indicating improvement) with a small effect size post-intervention (change score – 2.87±10.47, CI95=[-12.56; 6.82], d=-0.27) and a moderate effect size at follow-up (change score – 3.37±6.50, CI95=[-10.18; 3.45], d=-0.52). However, self-esteem and independence decreased (indicating worsening) in the exercise group, demonstrating moderate to large effect sizes post-intervention (self-esteem: change score –3.87±6.38, CI95=[-9.77; 2.03], d=-0.61; independence: change score –2.14±3.53, CI95=[-5.41; 1.12], d=-0.61) and at 6-month follow-up (self-esteem: change score –5.73±6.76, CI95=[-12.83; 1.36], d=-0.85; independence: change score –2.33±1.51, CI95=[-3.91; –0.75], d=-1.55).

### 3.2 Brain imaging outcomes

For resting-state fMRI, there were no connectivity changes found in either group.

For the task-based fMRI, we found high heterogeneity in the results in the remotely delivered CMR group (n=6) and thus there were no significant group changes found. After the remotely delivered exercise (n=5), increased activity was found in the posterior parietal cortex for left leg movement imagery (left superior parietal lobe, bilateral activation in the supramarginal gyrus and angular gyrus), and lateral occipital complex (LOC), which is responsible for visual body perception (superior part) and spatial awareness (inferior part of the LOC) (Z = 3.27, p < 0.001 FWE-corrected, 929 voxels). For imaging sensation in the left leg, increased activation was found in the right precentral and postcentral gyrus, and superior temporal gyrus (Z = 3.14, p < 0.001 FWE-corrected, 1869 voxels), as well as a second cluster spanning the right LOC (superior and inferior divisions), and angular gyrus (Z = 3.56, p < 0.001 FWE-corrected, 1172 voxels). Surprisingly, the whole-body kinesthetic imagery only revealed increased activation in the left hemisphere, specifically in the precentral and postcentral gyrus, superior and middle frontal gyri, supramarginal gyrus, angular gyrus, superior parietal lobe, and LOC (superior division) (Z = 3.55, p < 0.001 FWE-corrected, 6697 voxels). Finally, the right toe sensation task elicited increased activation in a left-hemisphere cluster encompassing the left supramarginal gyrus, angular gyrus, LOC (superior and inferior divisions), and precentral and postcentral gyrus (Z = 3.55, p < 0.001 FWE-corrected, 6697 voxels; **Figs. 4A-D**, **Table 3**).

**Fig. 4.**
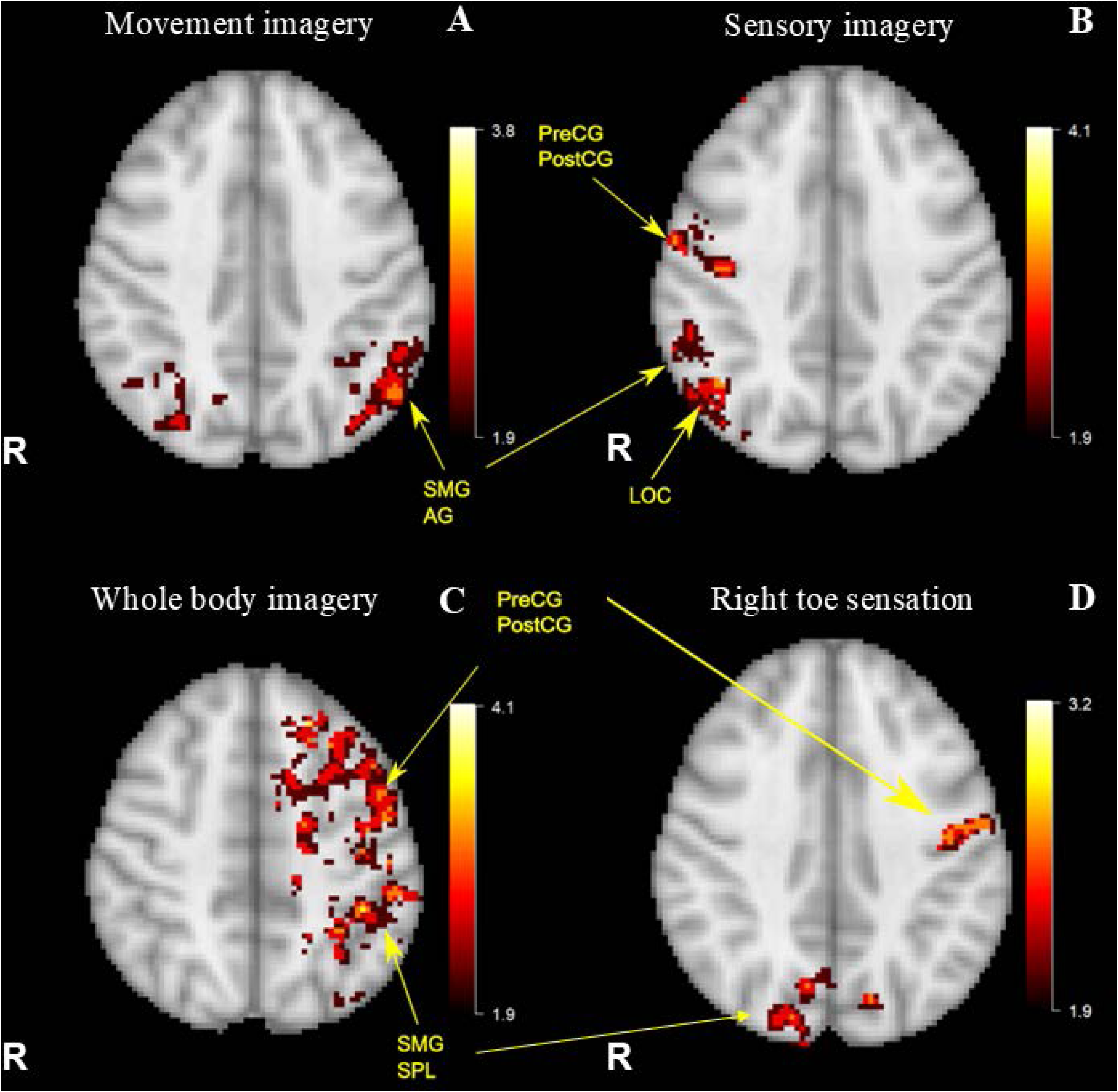
Pre-post task-based fMRI changes in brain activity in the remotely delivered exercise group. (**A**) Movement imagery (left leg): Increased activation post-exercise in the left supramarginal gyrus (SMG), angular gyrus (AG), and lateral occipital complex (LOC, superior and inferior divisions). **(B)** Sensory imagery (left leg): Increased activation post-exercise in the right precentral (PreCG) and postcentral gyrus (PostCG), lateral occipital complex (LOC, superior and inferior divisions), and angular gyrus (AG). **(C)** Whole-body imagery: Left hemisphere activation is increased after exercise, specifically in the precentral (PreCG) and postcentral gyrus (PostCG), supramarginal gyrus (SMG), angular gyrus (AG), and the superior parietal lobe (SPL), and the lateral occipital complex (superior division). **(D)** Right toe sensation: Left supramarginal gyrus (SMG), angular gyrus (AG), lateral occipital complex (LOC, superior and inferior divisions), precentral (PreCG) and postcentral gyrus (PostCG) had increased activation after exercise.

## 4 Discussion

This pilot clinical trial provides preliminary evidence that both Cognitive Multisensory Rehabilitation (CMR) and an active exercise control can be safely and feasibly delivered remotely to adults with spinal cord injury (SCI). The main findings were that, while both interventions yielded benefits, the remotely delivered CMR group exhibited large, sustained effect sizes for objective sensorimotor recovery (AIS and NRS), and specific neuropathic pain characteristics (DN4).

Although reductions in highest neuropathic pain across the whole cohort did not meet the established MCID for NPRS (averaging 1.25 points post-intervention and 0.86 points at 6-month follow-up), CMR successfully drove a large, meaningful reduction in qualitative neuropathic pain signatures, as evidenced by a large effect size on the DN4 post-intervention (d=-0.90) that was sustained at the 6-month follow-up (d=-0.71). Instead, the remotely delivered exercise group demonstrated smaller effect sizes for sensorimotor improvements and only a 0.57-point reduction for highest neuropathic pain (NPRS) immediately after exercise, and no changes for neuropathic pain characteristics (DN4) immediately after exercise and a slight increase in those characteristics at 6-month follow-up.

These findings suggest that while generalized exercise provides broad systemic benefits, directly targeting the brain’s internal body map via CMR may be necessary to drive specific, complex sensorimotor and neuropathic recovery in SCI. Contrary to our hypotheses and prior in-person studies, task-based fMRI revealed no significant group-level neural changes in the CMR cohort due to high inter-subject heterogeneity, whereas the exercise group showed increased activation in visuospatial and sensorimotor cortical regions.

The recovery seen in the CMR cohort aligns with our previous in-person CMR studies in adults with SCI, confirming the role of MBR for functional recovery.(22,23) The results observed after CMR support our premise that actively restoring MBR can drive functional recovery in chronic SCI. Because CMR specifically targets the brain’s internal body map through multisensory discrimination and kinesthetic imagery, participants may learn to better process residual sensory feedback, creating the necessary neural conditions to re-engage paralyzed or paretic limbs.(22,23) CMR may help facilitate the reciprocal interaction between the sensory and motor systems—a key prerequisite for skilled movement and sensorimotor relearning.(69) In contrast, the exercise group demonstrated smaller effect sizes for these same measures. In line with these objective measures, participants in the CMR group demonstrated large effect sizes for self-reported basic mobility, self-care, fine motor function, and ambulation on the (SCI-FI/AT) compared to moderate improvements in the exercise group. Both groups showed large effect sizes with regards to their self-identified functional goal attainment and in general, we received feedback that participants were satisfied and found benefit from their respective programs.

The reduction of overall pain intensity in the active exercise cohort aligns with the scant previous existing literature demonstrating that submaximal exercise can reduce neuropathic pain by 0.85 points.(70) While general exercise may adequately modulate broad pain intensities or specific musculoskeletal nociceptive pain, it often fails to consistently resolve central neuropathic pain.

Indeed, a comprehensive Cochrane review of non-pharmacological interventions for chronic pain in SCI concluded that standard modalities—including generalized exercise programs—frequently yield insufficient or inconsistent efficacy for deep neuropathic relief.(71)

Central neuropathic pain post-SCI is widely understood to be driven by maladaptive cortical reorganization, particularly within the primary somatosensory cortex, insula, and thalamus, resulting from sensory deafferentation.(72) Aside from reducing neuropathic pain intensity,(22,23) CMR also helps alter the qualitative, neuropathic signatures of the pain, yielding a distinct, centrally-mediated therapeutic mechanism that may address the limitations of standard physical exercise.

CMR also resulted in small-to-moderate reductions in spasticity frequency and severity at the two timepoints, whereas only spasm severity decreased with small to moderate effect in the exercise group. Notably regarding sleep quality, while both groups demonstrated initial improvements post-intervention, sustained moderate improvements in sleep were found at 6-months follow-up after CMR whereas the exercise group regressed back to baseline levels.

A surprising finding was the lack of group-level task-based fMRI changes in the CMR cohort. This contrasts with our previous in-person CMR trials, which demonstrated significant activation increases within the parietal operculum, insula, and posterior parietal cortex. This discrepancy was primarily driven by high inter-subject heterogeneity. Several factors may account for this variance. First, transitioning to a remote delivery modality may introduce variability in how participants engage with kinesthetic imagery and multisensory discrimination tasks without the hands-on physical guidance of an in-person therapist. Second, the broad clinical inclusion criteria regarding SCI severity, neurological level of injury, and baseline pain levels likely contributed to diverse baseline neural architectures and varied functional reorganization strategies across participants. Third, given the pilot sample sizes, these preliminary neuroimaging results must be interpreted with caution.

Interestingly, the remote exercise group demonstrated post-intervention brain activation increases in the left supramarginal and angular gyri (components of the posterior parietal cortex) during movement imagery, as well as increased right-hemisphere precentral and postcentral activation during sensory imagery. Notably, the lateral occipital complex (LOC) exhibited extensive activation clusters across all four evaluated tasks, encompassing movement imagery, sensory imagery, whole-body imagery, and right toe sensation. This LOC activation post-exercise suggests that engaging in standard-of-care remote physical exercise may promote neural recruitment in regions associated with somatosensory processing and visuospatial awareness, albeit potentially through a distinct mechanistic pathway compared to CMR. Indeed, in our prior studies, the LOC was not activated; this absence may stem from the fact that the LOC is primarily a visual processing area, and CMR exercises are predominantly performed with eyes closed during the learning phase to improve MBR. Following this, at the conclusion of each session, these improved MBR are then actively integrated into functional tasks, which are executed with eyes open.

Self-reported measures of interoception yielded intriguing mixed results, tentatively suggesting that both interventions may influence the brain’s internal body map, albeit potentially through different mechanisms. On the MAIA-2, the remote CMR group demonstrated moderate improvements in the “noticing” subdomain post-intervention, and a large, delayed improvement in the “not worrying” subdomain by the 6-month follow-up. This could indicate that CMR may help participants reduce the emotional distress associated with abnormal body sensations over time as they rebuild a more accurate MBR. Interestingly, the remote exercise group also demonstrated improvements in interoceptive awareness, yielding large effect sizes for “body listening” post-intervention that further increased at the 6-month follow-up, alongside large improvements in “trusting” their body post-intervention, evolving to moderate improvements when 6-month follow-up was compared to baseline. Together, these preliminary data suggest that while CMR may specifically help mitigate worry regarding dysregulated bodily sensations, standard vigorous exercise might also be effective at promoting active bodily listening and trust in those body signals.

Several limitations are worth noting. Most notably, the small sample size (n=15) limits the strength of interpretation of the behavioral and brain imaging results. Furthermore, the transition to a remote delivery model introduces variability; for instance, the reliance on caregivers for physical assistance during remote CMR introduces a potential variable in the consistency of the multisensory feedback provided, which was not heavily controlled for. Future, fully powered multi-site randomized controlled trials are urgently needed to confirm these preliminary findings. While our pilot trials have had a fairly equal distribution of adults with paraplegia vs tetraplegia, larger trials would allow us to perform sub-analyses by injury level (paraplegia vs tetraplegia) or other SCI-related variables. Next, all participants were white, non-Hispanic participants, with 13% veteran representation, 47% living in rural locations, and 20% reaching about 50% above the poverty level, reflecting moderate financial self-sufficiency. While we reached some metrics of health disparities, and almost 50% of those living rurally, more research is needed on those with SCI of racial and ethnic minorities.

In sum, a critical contribution of this study is the demonstration that both CMR and active exercise can be safely and effectively delivered remotely to adults with SCI. Transportation barriers are a well-documented hindrance to continuous rehabilitation for individuals with SCI, particularly those living in rural or underserved areas. The ability to achieve large effect sizes in sensorimotor functional recovery via secure videoconferencing provides a highly accessible, low-burden therapeutic avenue. For CMR specifically, having a trained caregiver assist the participant under the remote guidance of a certified therapist proved to be a viable model for delivering this complex, mind-body intervention.

## 6 Author Contributions

Every author contributed to the study: Conceptualization (AVdW); Funding acquisition (AVdW); Data curation (AVdW, SC, WD); Formal analysis and data interpretation (AVdW, LZ, TH, KL, BM); Remotely delivered Cognitive Multisensory Rehabilitation intervention (SB); Remotely delivered Exercise (RLL, ADR), Oversight of exercise program (AAH), Scheduling of the exercise program (SJL); Recruitment (AVdW); Data collection (AVdW, SC, WD, BM); General supervision (AVdW); Validation (AVdW, SC, WD, LZ); Visualization (AVdW, LZ); PM&R screening for exercise readiness (RN); SCI study doctor (LM); Writing – original draft (AVdW); Writing – review & editing (AVdW, AAH, SC, SB, RLL, ADR, SJL, WD, LZ, TH, BM, RN, LM, KL). We confirm that more than one author has directly accessed and verified the underlying data reported in the manuscript (AVdW, SC, LZ).

## 8 Acknowledgments

We thank Dr. Marco Rigoni and Dr. Marina Zernitz, Director and former Director of the Centro Studi di Riabilitazione Neurocognitiva – Villa Miari (Study Center for Cognitive Multisensory Rehabilitation), Italy, for her consultancy with the content of the CMR training protocol. We would like to thank the MRI technicians Wendy Elvendahl and Matthew White for their help with data acquisition. We thank Jena Blackwood for her assistance with data verification. We would like to extend our profound thanks to Marc Noël for the critical review of the manuscript.

## 9 Data Availability Statement

The dataset generated for this study can be found in the Dryad repository, DOI: 10.5061/dryad.vt4b8gv7f

## List of acronyms

AIS: ASIA Impairment Scale
ANS: Autonomic Standards Assessment form
BARQ-R: Revised Body Awareness Rating Questionnaire
CHART-SF: Craig Handicap Assessment and Reporting Technique-Short Form
CMR: Cognitive Multisensory Rehabilitation
DN4: Douleur Neuropathique 4
FAS: Functionality Appreciation Scale
fMRI: functional Magnetic Resonance Imaging
ISNCSCI: International Standards for Neurological Classification of Spinal Cord Injury
KVIQ: Kinesthetic and Visual Imagery Questionnaire
LOC: Lateral occipital complex
MAIA-2: Multidimensional Assessment Interoceptive Awareness – version 2
MBR: Mental Body Representations
MCID: Minimum Clinically Important Difference
MSES: Moorong Self-Efficacy Scale
NPRS: Numeric Pain Rating Scale
PAS: Postural Awareness Scale
PHQ-9: Patient Health Questionnaire
PSFS: Patient-Specific Functional Scale
PSQI: Pittsburgh Sleep Quality Index
REDCap: Research Electronic Data Capture
RPE: Borg Rating of Perceived Exertion
SAI/TAI: State Trait Anxiety Inventory
SCI: Spinal Cord Injury
SCI-FI/AT: Spinal Cord Injury Functional Index/Assistive Technology
SCI-QOL: Spinal Cord Injury Quality of Life measurement system
TSK: Tampa Scale for Kinesiophobia

## Data Sheet 1. Example of remotely delivered CMR exercises with two participants with SCI

### Participant 1

This woman in her 30’s sustained a C1 and T4 fracture resulting in a SCI (AIS D) from a car accident 16 years ago. At baseline, right upper limb strength was 4/5 (manual muscle testing) for wrist extensors and little finger abductor, and 5/5 for elbow flexors, elbow extensors, and finger flexors. Left upper limb strength was 5/5 for wrist extensors, 4/5 for elbow flexors and elbow extensors and 0/5 for finger flexors and little finger abductor. Leg strength was reduced to 4/5 for hip flexors on both sides, and to 3/5 for ankle dorsiflexors on the left side. Touch and pinprick sensation were reduced at T10 level and below.

After CMR and at the 6-month follow-up, strength was normal (5/5) on the right side and left upper limb strength improved to 5/5 for elbow flexors, 4/5 for finger flexors, and 3/5 for little finger abduction. Sensation for touch and pinprick fully returned throughout the body.

Functionally, core strength was impaired: she could only maintain balance while sitting on the treatment table when reaching forward or laterally less than 5 inches. Sit up, reversed sit-up, and trunk extension required considerable effort. The latter could only be performed with arms hanging down, not with hands behind the head. She was able to raise a 1-pound dumbbell in overhead press, but she could only lift her left elbow above shoulder level without a dumbbell. Shoulder flexion and door pull were decreased on the right side, and the left arm function was more severely affected, e.g., she could not grasp an empty can, remove the lid from a yoghurt container, or pick up a key with a pinch grip. She could initiate weightbearing on the legs (less than 50% upright) with inappropriate kinematics, maintain standing for at least one minute, and shift her body laterally with improper body mechanics.

After CMR and at 6-months follow-up, core strength gradually improved, enabling her to maintain balance with arms raised to the side and reach beyond 10 inches. She could perform a reverse sit-up to halfway, rotate her trunk bilaterally in this position and return to sitting without touching the mat (the maximum score for reversed sit-up). Trunk extension could be done easily and with hands behind the head. Right arm strength fully recovered (maximum score on all items), and the left arm improved in shoulder flexion to 90 degrees, grasp (moving empty can from mouth to table) and pinch grip (grasping and turning a key by supinating and pronating the forearm with improper kinematics). She could also open the lid of the yogurt container with her left hand (while holding the container with her right hand) and began reaching into the container to pick up a bolt. She was able to sit on the edge of the treatment table with feet on the floor and stand up with inappropriate kinematics but without using her arms as counterbalance. She maintained standing with proper body mechanisms and could keep her balance when pushed, with the body moving no more than 2 inches. She was able to take repetitive steps with appropriate body mechanics.

At baseline she experienced neuropathic pain on the right foot stimulated by touching something, right arm, left shoulder and neck (Numeric Pain Rating Scale for highest neuropathic pain: 6/10). At baseline, during transfers, her left foot lost contact with the floor at the heel, accompanied by spasms. While standing, she was unable to properly shift body weight over the left foot. Transfers were accomplished mainly using the right side of the body.

She enjoyed drawing and illustrated her perception of the altered MBR. This picture was drawn 3 weeks after receiving CMR, on the 9^th^ therapy session.

**The goal of the exercise is for the person, with eyes closed, to identify where the carpet is placed under the left foot. The carpet could be placed under the heel, midfoot, ball of the foot, or toes**.

**The therapist aims to restore the participant’s perception of the foot by remapping its length, shape, and dimension, as well as interpreting and integrating localization of touch (through the carpet under the foot).** Increased awareness of the foot’s dimensions will help the participant shift body weight over *the total surface of the foot* and maintain contact with the floor while standing. In addition, this awareness serves as preparatory skill for shifting body weight onto the left foot while standing.

To facilitate solving the exercise, the therapist allowed her to see and feel the position of the carpet under her right foot, providing a ‘model’ for the comparison with the sensation in her left foot. This approach also helps reinforce the brain’s memory of the right foot’s length, shape and dimensions, supporting the restoration of this awareness in the left foot.

### Participant 2

This man in his 40’s sustained a SCI at the level of L2 (AIS A) from a car accident 12 years ago. At baseline, there was no muscle contraction below the knee, and touch and pin prick sensation in the lower legs and feet was reduced or absent.

After CMR, trace muscle activation returned in the right ankle dorsiflexors, and in the ankle plantar flexors on both sides. At the 6-month follow-up, muscle traces of ankle plantar and dorsiflexors were present in both legs, along with a muscle trace of long toe extensors in the left foot. Touch and pin prick sensation levels remained largely unchanged (changes of 0-2 points).

Core strength improved from being able to perform a reverse sit-up (only 1/4^th^ of the way) and initiate a sit up at baseline, to being able to complete both tasks fully, post-CMR and at the 6-month follow-up. As he had paraplegia, he already achieved the maximum score for arm function at baseline. Finally, at baseline, the participant could initiate standing by shifting his body weight over his toes (less than 50% upright) using inappropriate kinematics. After CMR and at 6-month follow-up, he was able to stand up from a seated position with hip flexors at 80 degrees and stand without assistance.

In this exercise, the feet are positioned on a homemade balance board. The top wooden panel can be inclined so that the heels are lower than the toes (as shown); placed directly on top of the underlying panel (aligning heels and toes); or positioned farther forward so the toes are lower than the heels. The underlying panel can also be set on one side, making one foot higher than the other.

**The goal of the exercise is for the person, with eyes closed, to identify the position of their feet. The therapist aims to restore the participant’s recognition of the position of the heels relative to the toes and the spatial relationship between the legs, given the loss of sensation and MBR in both lower legs and feet.** To achieve this, the exercise is designed so the participant focuses on kinesthetic information (sensations) in the ankles and the position of the feet in space. The brain must integrate multiple types of sensory information from the legs and the feet to restore MBR.

**The therapist uses specific questions to help the participant restore their MBR.** The therapist asks: “If you were hiking, based on this sensation, where would the mountain be relative to you? In front of you, behind you, to your left or to your right? Or are you walking on flat terrain?”

In this way, the therapist helps the patient access a pre-injury image. The content of the imagery will be adapted to the participant’s prior experiences. For example, this participant enjoyed the outdoors). This pre-injury memory and imagery aid the participant in better identifying the current sensation in the lower legs by referencing a pre-injury image of the whole body).

This exercise will also help the participant regain the role of the feet as a ‘base’, improving the organization of the base of support ─ an essential factor for standing up and maintaining balance while standing.

## Data Sheet 2. Example of remotely delivered exercises with two participants with SCI

### Participant 1

This man in his 30’s with paraplegia. He sustained a T10 injury (AIS A) from an iatrogenic injury at birth. He had a hip and spine fusion. At baseline, he had normal arm strength (5/5 on all items for AIS + maximum score on NRS) and 0/5 for leg strength on AIS and minimum score on NRS, which remained at post-intervention and 6-month follow-up. AIS Touch and pinprick surprisingly got worse over time (touch (max 54): 45; 41.5; 39 for average across the two limbs at the 3 timepoints; and pinprick (max 54): 43, 39.5, 38.5 points. Core strength (NRS) improved by one point each time. Subjectively, his partner and him both noticed an improvement in balance.

An example of an exercise that the participant performed after 1 month of remotely delivered exercise was the following: The participant was seated in a manual wheelchair in his home office. He independently anchored the resistance band overhead to a closed door using commercial resistance band anchor loop and grabber.

Holding the resistance bands in both hands, he began with shoulders in flexion and elbows fully extended out in front of body. The participant pulled the hands down towards side of manual wheelchair keeping the elbows straight in extension and wrists neutral engaging in shoulder extension through range of motion until arms reach side of trunk. Then, keeping elbows straight, the participant returned to the starting position and repeated these movements 10 repetitions per set of 3.

The exercise physiologist gave cues to maintain upright seated posture and engage his core throughout the activity, and to refrain from shrugging the shoulders and arching the back.

### Participant 2

This man in his 60’s sustained a SCI (AIS D) after a bicycle accident, by going over the handle bikes and stretched and hyper extending the neck upon landing, resulting in bruised (but not fractured) C3 vertebrae. At baseline, his right arm strength was 5/5 for elbow flexors and extensors, 4/5 for wrist extensors and finger flexors, and 0/5 for the abductor of the little finger. In his left arm, elbow flexors, wrist extensors, and finger flexors were 5/5, elbow extensors were 4/5 and the abductor of the little finger was 0/5 on the AIS test. This upper limb strength stayed the same through the study (for AIS and NRS). This leg strength (AIS) was normal on the left side (5/5 for all muscles) and 5/5 on the right side, except for the long toe extensors (4/5 at baseline and 6-month follow-up, 5/5 right after the exercise intervention). Touch and pinprick sensation (AIS) were normal in the whole body at baseline and remained so after exercise and at 6-month follow-up. Core strength (36 points at baseline on NRS) improved to 38 after the intervention and back to 37 at 6-month follow-up. The change score happened for trunk extension. Walking improved from taking repetitive steps with appropriate kinematics at baseline to being able to descend an incline after the exercise intervention and at 6-month follow-up.

Here is an example of an exercise that the participant performed after 12 sessions (one month) of remotely delivered exercise: The participant began seated upright in his power wheelchair with feet flat on the ground. The participant flexed forward at the trunk, initiating weight bearing evenly through feet; then extended at knees and hips to achieve full upright standing position without the use of his hands. The participant then returned to the seated position and repeated these movements 20 times per set of 2; completed in sequence with other high-repetition movements (e.g. marching, trunk rotations) as part of an interval training for the aerobic activity portion of the remotely delivered exercise program. The exercise physiologist provided verbal cues to encourage participant to exercise at a moderate-to-vigorous intensity while maintaining continuous movement throughout the activity.

**Supplementary Table 1.**
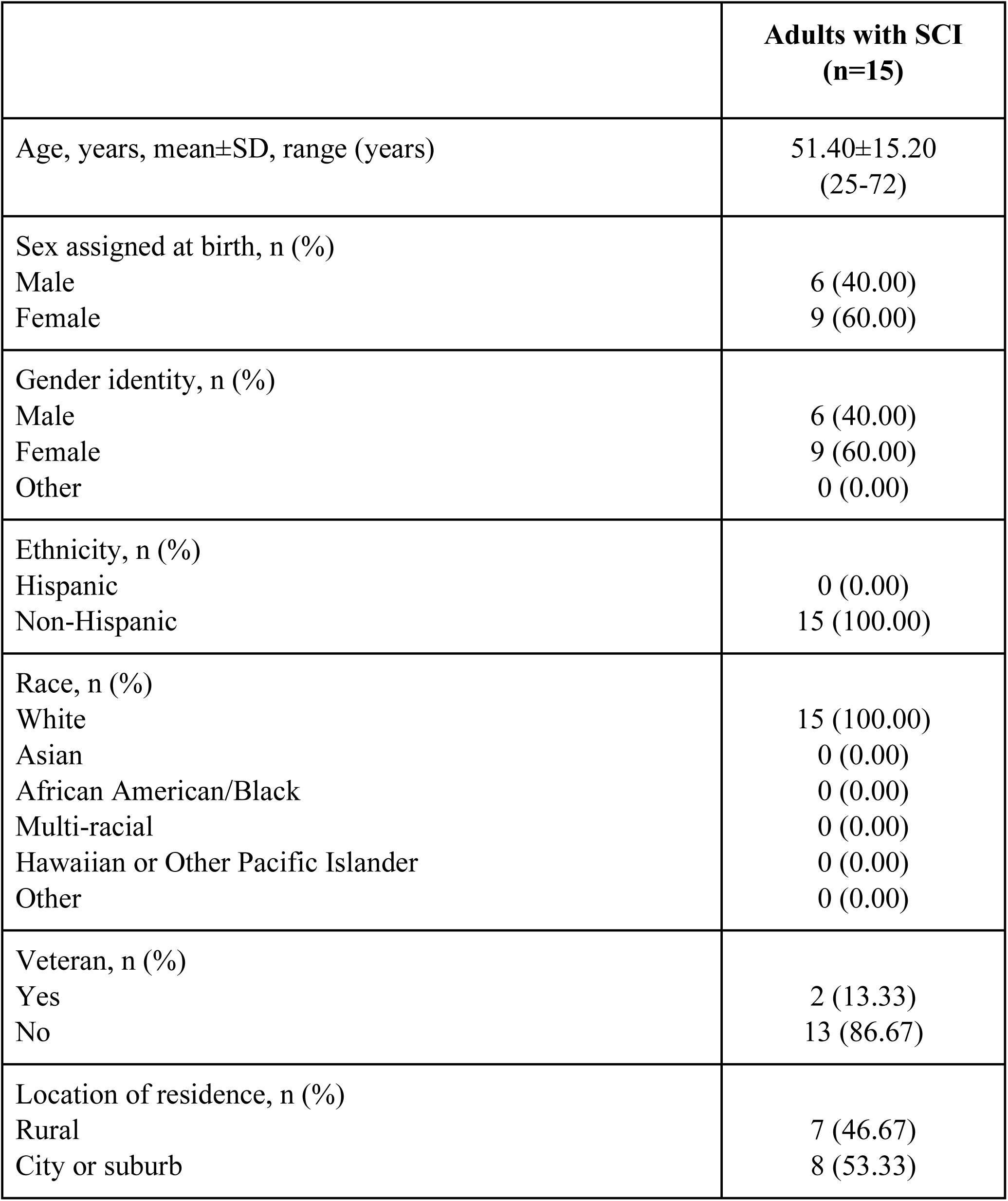
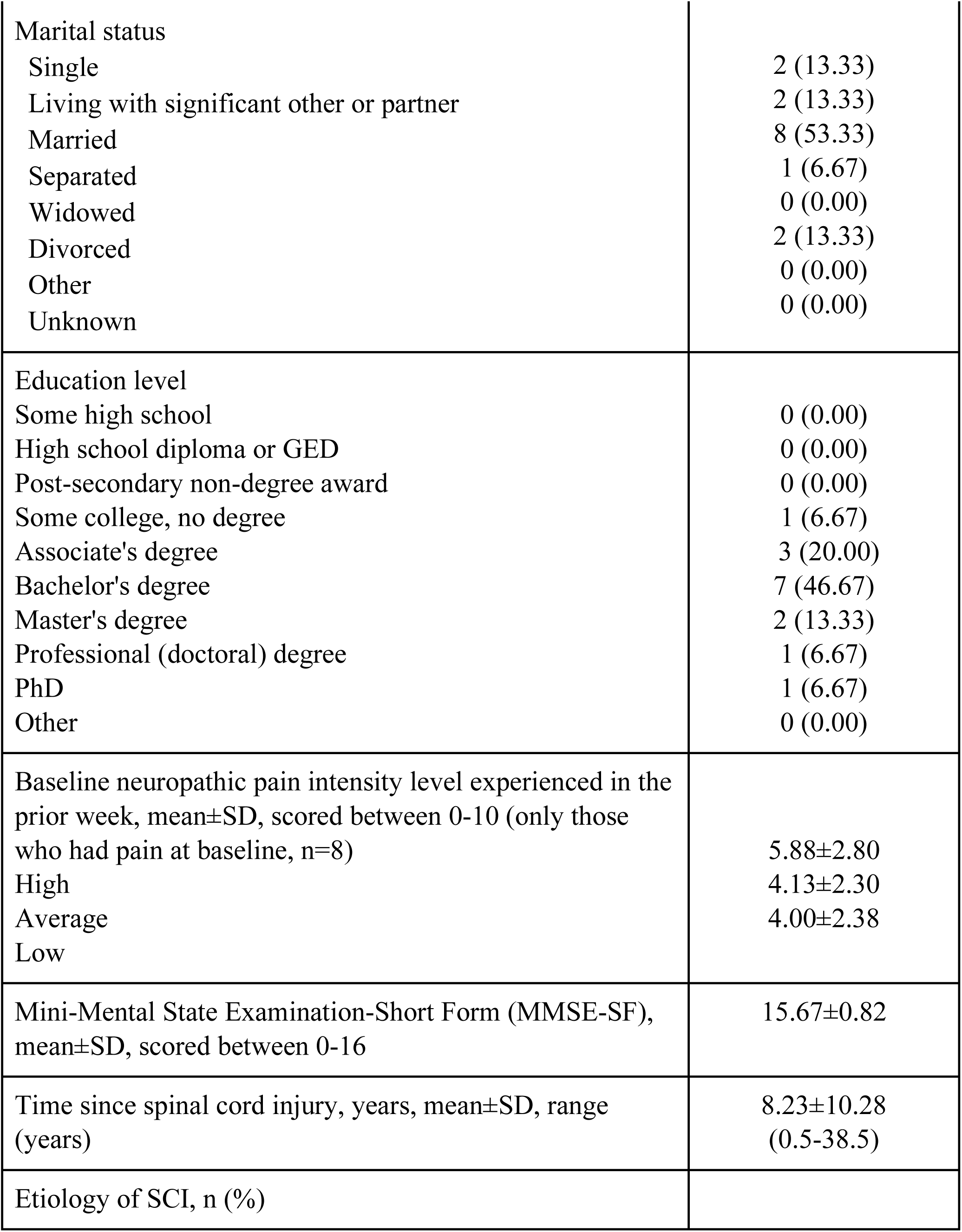
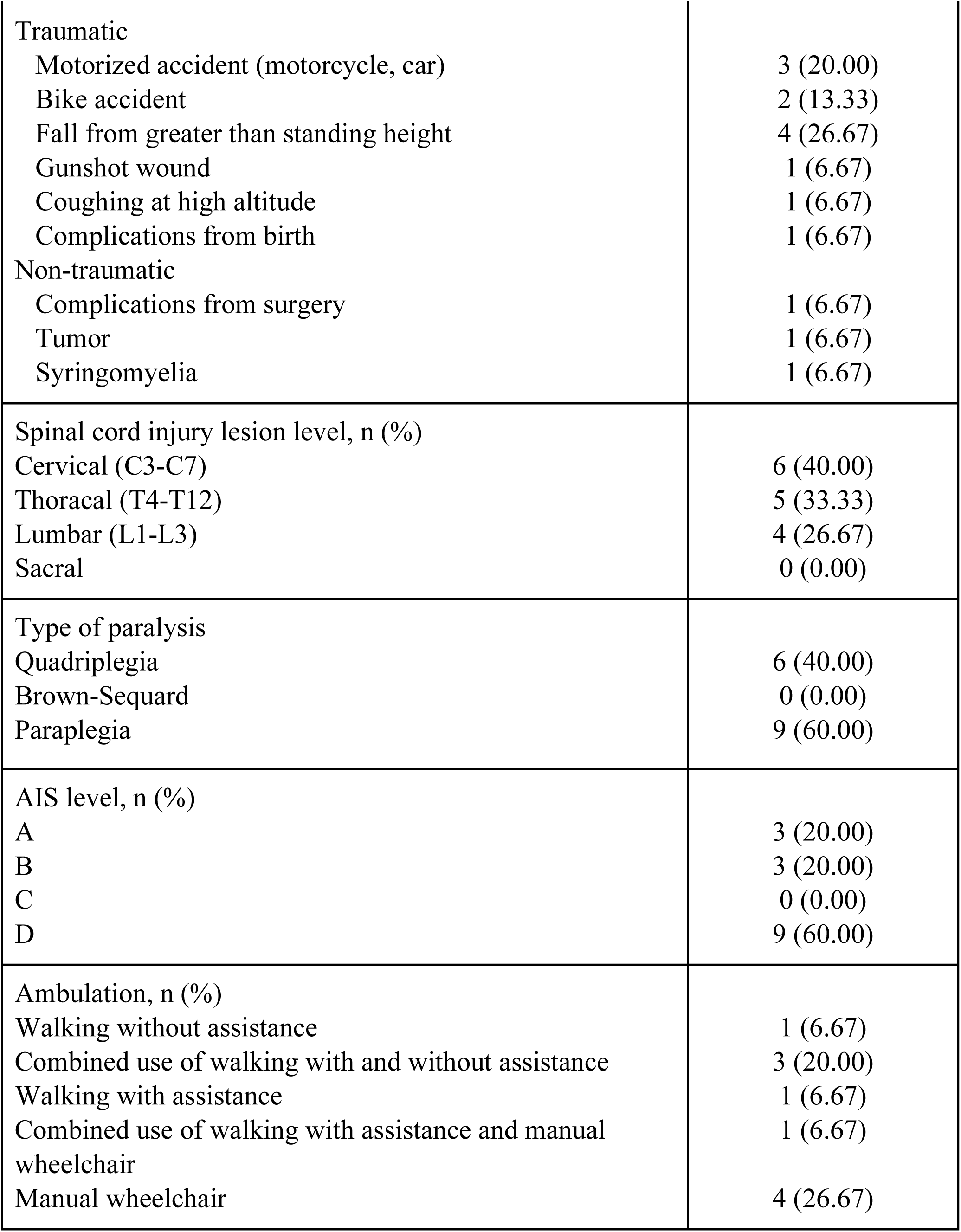
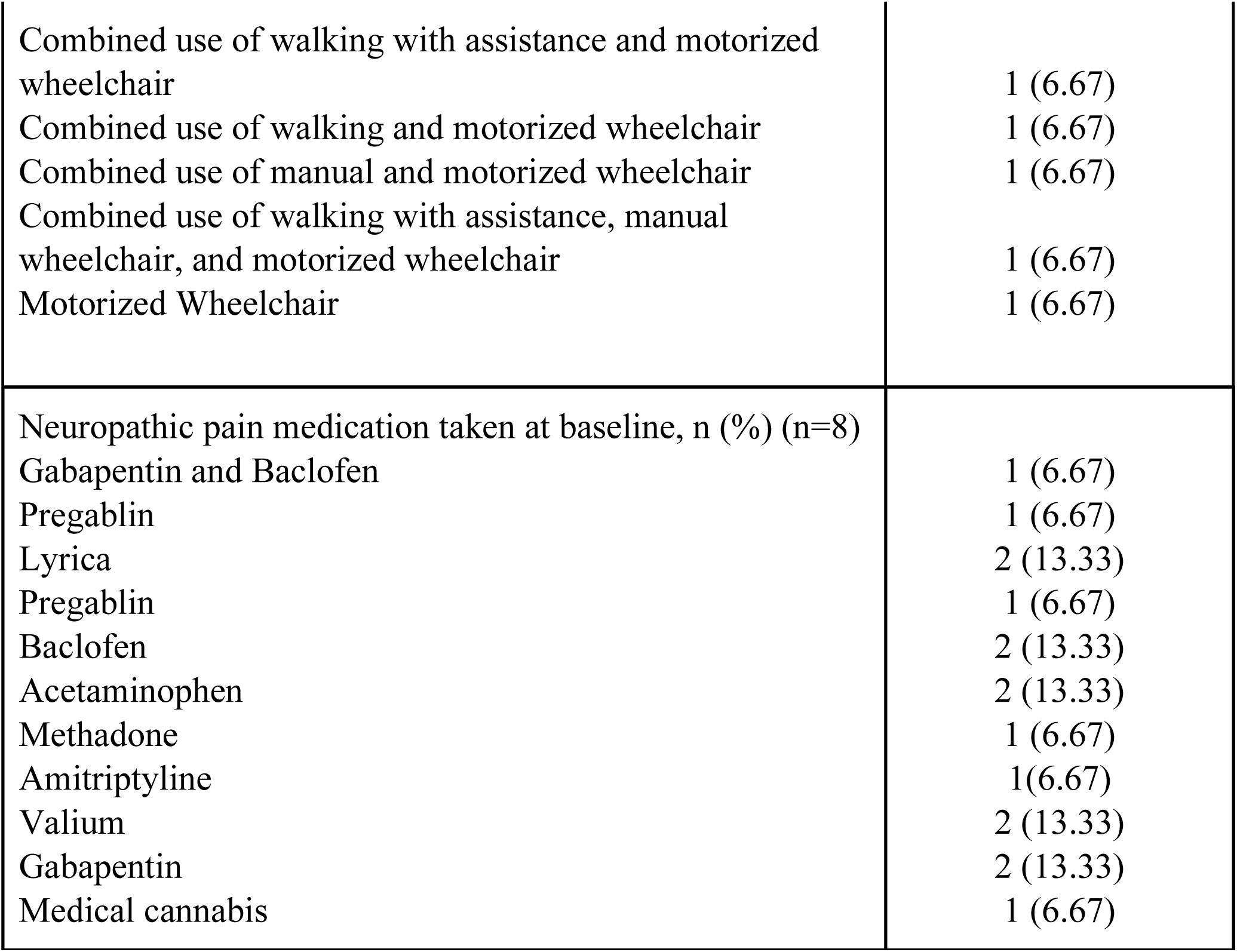
Demographic and clinical characteristics of adults with spinal cord injury.

**Supplementary Table 2.** Primary and secondary outcome measures at three time points.

| Outcome measures | Time point 1<br>Baseline<br>(mean±SD) |  | Time point 2<br>Post-intervention<br>(mean±SD) |  | Time point 3<br>6MFU<br>(mean±SD) |  | Effect size<br>Pre – Post |  | Effect size<br>Pre – 6MFU |  |
| --- | --- | --- | --- | --- | --- | --- | --- | --- | --- | --- |
|  | Remote<br>CMR | Remote<br>Exercise | Remote<br>CMR | Remote<br>Exercise | Remote<br>CMR | Remote<br>Exercise | Remote<br>CMR | Remote<br>Exercise | Remote<br>CMR | Remote<br>Exercise |
| n | (n=8) | (n=7) | (n=8) | (n=7) | (n=7) | (n=7) |  |  |  |  |
| **Highest NPRS** | 3.00±4.28 | 3.29±3.04 | 1.75±2.43 | 2.71±3.04 | 1.14±1.95 | 2.86±3.08 | -0.61 | -0.73 | -0.55 | -0.55 |
| **Average NPRS** | 1.75±2.87 | 2.14±2.12 | 0.88±1.36 | 1.43±1.72 | 0.57±1.13 | 1.86±1.95 | -0.48 | -0.57 | -0.38 | -0.38 |
| **Lowest NPRS** | 1.25±2.82 | 0.71±0.95 | 0.50±0.93 | 0.57±0.98 | 0.29±0.76 | 0.71±0.95 | -0.35 | -0.38 | NaN | NaN |
| **DN4** | 4.38±3.81 | 4.29±3.15 | 3.25±3.20 | 4.29±3.15 | 2.86±3.44 | 4.43±3.21 | -0.90 | NaN | -0.71 | 0.38 |
| n | (n=8) | (n=7) | (n=8) | (n=7) | (n=7) | (n=7) |  |  |  |  |
| **AIS exam** |  |  |  |  |  |  |  |  |  |  |
| - sensory touch (Avg L, R) | 48.81±2.55 | 46.36±7.20 | 52.19±2.20 | 47.43±7.83 | 52.14±2.19 | 46.43±8.71 | 1.30 | 0.35 | 1.73 | 0.02 |
| - sensory pin (Avg L, R) | 47.13±4.37 | 44.21±7.48 | 51.81±2.56 | 45.21±8.18 | 51.21±3.60 | 44.50±7.77 | 1.21 | 0.36 | 1.09 | 0.09 |
| - lower limb motor function | 16.25±7.98 | 13.07±12.34 | 18.13±7.90 | 13.29±12.56 | 17.50±7.49 | 13.29±12.56 | 1.83 | 0.80 | 2.05 | 0.55 |
| **NRS** |  |  |  |  |  |  |  |  |  |  |
| - core | 36.13±8.18 | 27.86±14.75 | 43.88±8.10 | 28.29±15.61 | 44.14±7.20 | 28.43±15.86 | 1.48 | 0.31 | 1.72 | 0.32 |
| - upper limb | 63.36±15.07 | 57.50±18.45 | 67.38±8.71 | 59.07±16.95 | 67.29±8.38 | 58.71±18.87 | 0.69 | 0.74 | 0.58 | 0.47 |
| - lower limb | 16.00±9.01 | 17.86±11.38 | 21.88±11.46 | 19.57±12.15 | 21.00±11.02 | 20.29±12.98 | 1.25 | 0.83 | 1.38 | 0.88 |
| **PSFS** | 2.13±1.61 | 3.10±0.60 | 6.71±2.00 | 6.57±2.58 | 7.24±1.97 | 6.71±2.73 | 1.77 | 1.28 | 1.98 | 1.28 |
| n | (n=5) | (n=4) | (n=5) | (n=4) | (n=5) | (n=4) |  |  |  |  |
| **SCI-FI/AT paraplegia** |  |  |  |  |  |  |  |  |  |  |
| - basic mobility | 30.40±3.91 | 30.00±5.60 | 34.20±1.79 | 31.00±4.76 | 34.80±1.79 | 31.00±4.76 | 1.53 | 0.71 | 1.37 | 0.71 |
| - self-care | 30.80±4.21 | 34.75±1.89 | 33.80±3.90 | 35.00±1.41 | 34.20±3.03 | 34.50±2.38 | 1.10 | 0.50 | 1.36 | -0.50 |
| - fine motor function | 31.00±1.22 | 31.00±1.15 | 32.00±0.00 | 31.25±0.96 | 32.00±0.00 | 31.25±0.96 | 0.82 | 0.50 | 0.82 | 0.50 |
| - ambulation | 9.20±5.89 | 6.25±6.85 | 13.20±4.49 | 7.00±8.04 | 13.60±4.28 | 7.00±8.04 | 2.31 | 0.50 | 2.01 | 0.50 |
| n | (n=3) | (n=3) | (n=3) | (n=3) | (n=2) | (n=3) |  |  |  |  |
| **SCI-FI/AT quadriplegia** |  |  |  |  |  |  |  |  |  |  |
| - basic mobility | 18.33±2.89 | 26.00±11.79 | 29.67±5.86 | 26.00±11.79 | 29.00±5.66 | 25.67±12.34 | 1.41 | NaN | 1.59 | -0.58 |
| - self-care | 21.67±4.51 | 24.33±7.57 | 28.00±9.17 | 24.67±8.14 | 26.50±7.78 | 24.33±7.57 | 1.34 | 0.58 | 1.65 | NaN |
| - fine motor function | 19.67±4.16 | 20.33±3.79 | 24.67±7.51 | 21.00±4.36 | 24.50±7.78 | 20.33±3.79 | 1.39 | 0.58 | 1.84 | NaN |
| - ambulation | 8.33±3.51 | 12.33±10.79 | 15.00±3.00 | 12.67±11.02 | 13.50±2.12 | 12.67±11.02 | 2.18 | 0.58 | 1.65 | 0.58 |
| n | (n=8) | (n=7) | (n=8) | (n=7) | (n=7) | (n=7) |  |  |  |  |
| **Spasms** |  |  |  |  |  |  |  |  |  |  |
| - frequency | 1.13±1.46 | 1.14±0.69 | 0.63±0.74 | 1.14±0.69 | 0.57±0.79 | 1.14±0.69 | -0.47 | NaN | -0.38 | NaN |
| - severity | 1.00±1.20 | 1.57±0.79 | 0.63±0.74 | 1.43±0.79 | 0.43±0.53 | 1.29±0.76 | -0.50 | -0.38 | -0.55 | -0.59 |
| PSQI | 8.50±3.96 | 7.86±2.79 | 5.88±1.06 | 6.00±2.71 | 6.00±1.41 | 7.71±4.15 | -0.59 | -1.18 | -0.59 | -0.07 |
| Trait Anxiety Inventory | 38.38±6.84 | 34.71±6.63 | 36.38±7.01 | 35.43±8.89 | 35.43±9.50 | 33.57±8.16 | -0.47 | 0.21 | -0.45 | -0.39 |
| State Anxiety Inventory | 31.88±8.36 | 29.29±8.98 | 30.63±8.68 | 30.14±8.30 | 27.71±7.89 | 29.29±8.46 | -0.27 | 0.31 | -1.01 | 0.00 |
| PHQ-9 | 4.63±5.18 | 3.14±2.12 | 2.88±3.44 | 3.29±2.93 | 4.86±2.73 | 2.57±2.44 | -0.62 | 0.06 | -0.03 | -0.24 |
| TSK | 32.38±6.02 | 33.86±7.29 | 31.25±5.37 | 36.29±6.65 | 31.71±4.99 | 35.29±5.06 | -0.23 | 0.42 | 0.13 | 0.42 |
| ANS | 31.38±7.63 | 33.14±3.67 | 32.50±7.82 | 33.29±3.73 | 31.29±7.13 | 33.29±3.73 | 0.73 | 0.38 | 0.65 | 0.38 |
| MSES | 95.75±11.84 | 93.86±12.82 | 97.88±9.37 | 95.43±10.86 | 97.57±9.25 | 94.71±10.63 | 0.33 | 0.29 | 0.30 | 0.33 |
| FAS | 2.93±0.76 | 3.06±0.95 | 3.43±0.72 | 3.22±0.59 | 3.51±0.73 | 3.40±0.47 | 1.04 | 0.22 | 1.25 | 0.51 |
| PAS | 41.25±13.93 | 54.43±10.34 | 49.63±13.07 | 58.29±11.09 | 52.00±11.28 | 57.57±7.85 | 1.12 | 0.53 | 1.46 | 0.67 |
| BARQ-R | 15.75±5.18 | 15.14±4.78 | 14.25±3.99 | 13.57±4.65 | 14.86±4.45 | 14.00±6.66 | -0.49 | -1.39 | 0.05 | -0.33 |
| **MAIA-2** |  |  |  |  |  |  |  |  |  |  |
| - noticing | 3.59±0.72 | 3.50±0.91 | 4.16±0.69 | 4.18±0.43 | 3.75±0.69 | 4.14±0.64 | 0.77 | 0.78 | 0.05 | 0.98 |
| - not distracted | 1.44±1.04 | 2.48±0.94 | 1.31±0.94 | 2.12±1.53 | 1.74±1.30 | 2.67±1.17 | -0.14 | -0.44 | 0.20 | 0.12 |
| - not worrying | 3.03±0.80 | 3.26±0.70 | 3.28±0.79 | 3.06±0.75 | 3.71±0.60 | 2.69±0.75 | 0.39 | -0.32 | 1.06 | -0.83 |
| - attention regulation | 3.04±0.97 | 3.71±0.52 | 3.43±0.77 | 3.78±0.14 | 3.08±0.71 | 3.90±0.40 | 0.36 | 0.14 | -0.21 | 0.30 |
| - emotional awareness | 3.03±1.07 | 3.57±0.55 | 3.53±0.89 | 3.66±0.75 | 3.31±0.65 | 3.40±1.01 | 0.47 | 0.34 | 0.19 | 0.23 |
| - self-regulation | 3.06±1.02 | 3.29±0.42 | 3.16±1.11 | 3.50±0.46 | 3.07±0.45 | 3.32±0.75 | 0.24 | 0.39 | -0.31 | 0.05 |
| - body listening | 2.13±1.22 | 2.43±0.42 | 2.67±0.99 | 3.05±0.65 | 2.57±1.17 | 3.29±0.40 | 0.45 | 1.00 | 0.53 | 2.02 |
| - trusting | 2.92±1.63 | 3.38±0.76 | 3.33±1.22 | 3.95±0.59 | 3.10±1.36 | 4.00±0.38 | 0.37 | 1.01 | -0.42 | 0.75 |
| KVIQ | 48.13±2.36 | 47.14±2.85 | 49.00±1.77 | 49.86±0.38 | 49.57±0.79 | 49.86±0.38 | 0.48 | 1.06 | 0.52 | 1.06 |
| **CHART-SF** |  |  |  |  |  |  |  |  |  |  |
| - physical independence | 94.75±14.85 | 91.71±8.04 | 96.75±9.19 | 94.00±7.75 | 96.29±9.83 | 94.00±7.75 | 0.35 | 0.38 | 0.38 | 0.38 |
| - cognitive independence | 100.00±0.00 | 100.00±0.00 | 100.00±0.00 | 100.00±0.00 | 100.00±0.00 | 100.00±0.00 | NaN | NaN | NaN | NaN |
| - mobility | 81.50±26.80 | 85.86±18.40 | 93.63±10.24 | 87.29±16.12 | 94.00±9.00 | 87.29±16.12 | 0.53 | 0.38 | 0.68 | 0.38 |
| - occupation | 66.64±34.23 | 53.75±46.76 | 82.97±25.99 | 49.82±43.02 | 82.14±25.07 | 52.32±40.04 | 0.53 | -0.34 | 0.63 | -0.09 |
| - social interaction | 100.00±0.00 | 100.00±0.00 | 100.00±0.00 | 100.00±0.00 | 100.00±0.00 | 100.00±0.00 | NaN | NaN | NaN | NaN |
| - economic self-sufficiency | 94.23 ±9.91 | 90.01±17.44 | 94.23±9.91 | 90.01±17.44 | 96.64±7.04 | 90.01±17.44 | NaN | NaN | 0.38 | NaN |
|  | (n=8) | (n=7) | (n=8) | (n=7) | (n=7) | (n=6) |  |  |  |  |
| **SCI-QOL (T-scores)** |  |  |  |  |  |  |  |  |  |  |
| - positive affect | 53.75±5.49 | 55.26±5.21 | 54.56±6.61 | 55.33±5.52 | 55.94±7.78 | 54.50±6.06 | 0.17 | 0.08 | 0.49 | 0.05 |
| - resilience | 49.66±7.77 | 50.26±4.86 | 51.61±6.77 | 49.63±6.95 | 52.81±8.10 | 51.55±8.85 | 0.29 | -0.17 | 0.35 | 0.24 |
| - self-esteem | 32.38±7.90 | 33.61±6.79 | 31.91±7.13 | 29.74±9.57 | 32.37±9.52 | 28.87±8.15 | -0.08 | -0.61 | -0.04 | -0.85 |
| - ability to participate | 44.15±7.58 | 45.56±2.86 | 46.58±6.59 | 47.19±7.26 | 49.53±9.17 | 46.35±2.90 | 0.27 | 0.30 | 0.42 | 0.67 |
| - independence | 46.14±6.81 | 46.24±3.53 | 44.105±3.10 | 44.13±3.26 | 44.37±2.56 | 44.42±2.86 | -0.40 | -0.61 | 0.11 | -1.49 |
| - pain behavior | 51.89±9.51 | 50.76±3.28 | 51.48±8.62 | 47.89±9.31 | 50.84±9.07 | 46.37±9.04 | -0.04 | -0.27 | 0.00 | -0.52 |

**Legend:** ANS: Autonomic Standards Assessment Form; BARQ-R: Revised Body Awareness Rating Questionnaire; CHART-SF: Craig Handicap Assessment and Reporting Technique; DN4: Douleur Neuropathique (Neuropathic pain) 4; FAS: Functionality Appreciation Scale; ISNCSCI-AIS exam: International Standards for Neurological Classification of SCI American Spinal Cord Injury Association Impairment Scale; KVIQ: Kinesthetic and Visual Imagery Questionnaire; MAIA-2: Multidimensional Assessment of Interoceptive Awareness Version 2; MSES: Moorong Self-Efficacy Scale; NPRS: Numeric Pain Rating Scale (used to measure neuropathic pain in the prior week); NRS: Neuromuscular Recovery Scale; PAS: Postural Awareness Scale; PSFS: Patient Specific Functional Scale; SCI: spinal cord injury; PHQ-9: Patient Health Questionnaire-9; PSQI: Pittsburgh Sleep Quality Index; SCI-FI/AT: Spinal Cord Injury Functional Index/Assistive Technology Short Forms; SCI-QOL: Spinal Cord Injury-Quality of Life scales; TSK: Tampa Scale for Kinesiophobia. **Note:** Positive and negative effect size values indicate whether there was improvement or worsening, respectively, of outcome measures. One person in the adaptive fitness group withdrew before the follow-up testing, so averages and SD are calculated n-1 for this time point. Effect sizes pre-3MFU also reflect the reduced n. All *p*-values are adjusted using Benjamini-Hochberg False Discovery Rate (FDR) for multiple comparison corrections

**Supplementary Table 3.** Brain imaging outcomes for task-based fMRI in adults with SCI.

|  | Z<br>score | p-value<br>(FWE-corrected) | Cluster<br>extent<br>(voxels) | Hemi-<br>sphere | x<br>(mm) | y<br>(mm) | z<br>(mm) |  |
| --- | --- | --- | --- | --- | --- | --- | --- | --- |
| fMRI task: Imagining moving the left leg<br>Post-exercise > Baseline in adults with SCI |  |  |  |  |  |  |  |  |
| Cluster 1 | 3.27 | <0.001 | 929 | Left | -68 | -52 | 48 | **Superior parietal lobe**<br>**Supramarginal gyrus (bilateral)**<br>**Angular gyrus (bilateral)**<br>**Lateral occipital complex superior division**<br>**Lateral occipital complex, inferior division**<br>Precuneus |
| fMRI task: Imagining feeling the left leg<br>Post-exercise > Baseline in adults with SCI |  |  |  |  |  |  |  |  |
| Cluster 1 | 3.14 | <0.001 | 1869 | Right | 60 | -18 | -2 | **Precentral gyrus**<br>**Postcentral gyrus**<br>Superior temporal gyrus |
| Cluster 2 | 3.56 | <0.001 | 1172 | Right | 42 | -60 |  | **Lateral occipital complex superior division**<br>**Lateral occipital complex, inferior division**<br>**Angular gyrus**<br>Occipital fusiform gyrus |
| fMRI task: Whole-body imagery<br>Post-exercise > Baseline in adults with SCI |  |  |  |  |  |  |  |  |
| Cluster 1 | 3.55 | <0.001 | 6697 | Left | -14 | -32 | 60 | **Precentral Gyrus**<br>**Postcentral Gyrus**<br>Superior frontal gyrus<br>Middle frontal gyrus<br>**Superior Parietal Lobule**<br>**Supramarginal gyrus**<br>**Angular gyrus**<br>**Lateral occipital complex, superior division** |
| fMRI task: Right toe sensation<br>Post-CMR > Baseline in adults with SCI |  |  |  |  |  |  |  |  |
| Cluster 1 | 3.26 | <0.001 | 965 | Left | -64 | -40 | 22 | **Supramarginal gyrus**<br>**Angular gyrus**<br>**Lateral occipital complex superior division**<br>**Lateral occipital complex, inferior division**<br>Precuneus |
| Cluster 2 | 3.21 | 0.01 | 472 | Left | -52 | -10 | 34 | **Precentral Gyrus**<br>**Postcentral Gyrus**<br>Superior temporal gyrus, posterior<br>division |

## References

1. Leemhuis E, Esposito RM, De Gennaro L, Pazzaglia M. Go virtual to get real: Virtual reality as a resource for spinal cord treatment. Int J Environ Res Public Health. 2021 Feb 13;18(4):1819.

2. Kim HY, Lee HJ, Kim TL, Kim E, Ham D, Lee J, et al. Prevalence and Characteristics of Neuropathic Pain in Patients With Spinal Cord Injury Referred to a Rehabilitation Center. Ann Rehabil Med. 2020 Dec;44(6):438–49.

3. Leemhuis E, Giuffrida V, De Martino ML, Forte G, Pecchinenda A, De Gennaro L, et al. Rethinking the body in the brain after spinal cord injury. J Clin Med. 2022 Jan 13;11(2):388.

4. Vastano R, Costantini M, Widerstrom-Noga E. Maladaptive reorganization following SCI: The role of body representation and multisensory integration. Prog Neurobiol. 2022 Jan;208(102179):102179.

5. Allison DJ, Ahrens J, Mirkowski M, Mehta S, Loh E. The effect of neuropathic pain treatments on pain interference following spinal cord injury: A systematic review. J Spinal Cord Med. 2024 Jul 3;47(4):465–76.

6. Almeida C, Monteiro-Soares M, Fernandes Â. Should non-pharmacological and non-surgical interventions be used to manage neuropathic pain in adults with spinal cord injury? – A systematic review. J Pain. 2022 Sep;23(9):1510–29.

7. Van de Winckel A, De Patre D, Rigoni M, Fiecas M, Hendrickson TJ, Larson M, et al. Exploratory study of how Cognitive Multisensory Rehabilitation restores parietal operculum connectivity and improves upper limb movements in chronic stroke. Sci Rep. 2020 Nov 20;10(1):20278.

8. Van de Winckel A, Carpentier ST, Deng W, Bottale S, Zhang L, Hendrickson T, et al. Identifying Body Awareness-Related Brain Network Changes after Cognitive Multisensory Rehabilitation for Neuropathic Pain Relief in Adults with Spinal Cord Injury: Delayed Treatment arm Phase I Randomized Controlled Trial [Internet]. medRxiv. 2023 [cited 2023 Feb 10]. p. 2023.02.09.23285713. Available from: https://www.medrxiv.org/content/10.1101/2023.02.09.23285713v1

9. Van de Winckel A, Carpentier S, Deng W, Bottale S, Hendrickson T, Zhang L, et al. Identifying Body Awareness-Related Brain Network Changes After Cognitive Multisensory Rehabilitation for Neuropathic Pain Relief in Adults With Spinal Cord Injury: Protocol of a Phase I Randomized Controlled Trial. Top Spinal Cord Inj Rehabil. 2022 Nov 15;28(4):33–43.

10. Guterstam A, Björnsdotter M, Gentile G, Ehrsson HH. Posterior cingulate cortex integrates the senses of self-location and body ownership. Curr Biol. 2015 Jun;25(11):1416–25.

11. Osinski T, Martinez V, Bensmail D, Hatem S, Bouhassira D. Interplay between body schema, visuospatial perception and pain in patients with spinal cord injury. Eur J Pain. 2020 Aug;24(7):1400–10.

12. Mehling WE, Wrubel J, Daubenmier JJ, Price CJ, Kerr CE, Silow T. Body Awareness: a phenomenological inquiry into the common ground of mind-body therapies. Philos Ethics Humanit Med. 2011;6.

13. Donegan T, Ryan BE, Sanchez-Vives MV, Świdrak J. Altered bodily perceptions in chronic neuropathic pain conditions and implications for treatment using immersive virtual reality. Front Hum Neurosci. 2022 Nov 17;16:1024910.

14. Pazzaglia M, Haggard P, Scivoletto G, Molinari M, Lenggenhager B. Pain and somatic sensation are transiently normalized by illusory body ownership in a patient with spinal cord injury. Restor Neurol Neurosci. 2016 Apr 11;34(4):603–13.

15. de Vignemont F. Body schema and body image—Pros and cons. Neuropsychologia. 2010 Feb 1;48(3):669–80.

16. Berlucchi G, Aglioti SM. The body in the brain revisited. Exp Brain Res. 2010 Jan;200(1):25–35.

17. Pitron V, Alsmith A, de Vignemont F. How do the body schema and the body image interact? Conscious Cogn. 2018 Oct;65:352–8.

18. Corradi-Dell’Acqua C, Tomasino B, Fink GR. What is the position of an arm relative to the body? Neural correlates of body schema and body structural description. J Neurosci. 2009 Apr 1;29(13):4162–71.

19. Di Vita A, Boccia M, Palermo L, Guariglia C. To move or not to move, that is the question! Body schema and non-action oriented body representations: An fMRI meta-analytic study. Neurosci Biobehav Rev. 2016 Sep;68:37–46.

20. Eickhoff SB, Jbabdi S, Caspers S, Laird AR, Fox PT, Zilles K, et al. Anatomical and functional connectivity of cytoarchitectonic areas within the human parietal operculum. J Neurosci. 2010 May 5;30(18):6409–21.

21. Peyron R, Fauchon C. The posterior insular-opercular cortex: An access to the brain networks of thermosensory and nociceptive processes? Neurosci Lett. 2019 May 29;702:34–9.

22. Van de Winckel A, Carpentier ST, Deng W, Bottale S, Zhang L, Hendrickson TJ, et al. Preliminary efficacy of cognitive multisensory rehabilitation for neuropathic pain in chronic spinal cord injury: a phase I randomized controlled trial. Sci Rep [Internet]. 2026 Mar 20; Available from: 10.1038/s41598-026-44859-w

23. Van de Winckel A, Carpentier ST, Bottale S, Blackwood J, Deng W, Zhang L, et al. Greater sustained sensorimotor function recovery and neuropathic pain reduction with Cognitive Multisensory Rehabilitation compared to adaptive fitness in adults with spinal cord injury: a pilot clinical trial [Internet]. medRxiv. medRxiv; 2026. Available from: 10.64898/2026.03.26.26349257

24. Van de Winckel A, Wenderoth N, De Weerdt W, Sunaert S, Peeters R, Van Hecke W, et al. Frontoparietal involvement in passively guided shape and length discrimination: a comparison between subcortical stroke patients and healthy controls. Exp Brain Res. 2012 Jul;220(2):179–89.

25. Van de Winckel A, Sunaert S, Wenderoth N, Peeters R, Vanhecke P, Feys H, et al. Passive somatosensory discrimination tasks in healthy volunteers: Differential networks involved in familiar versus unfamiliar shape and length discrimination [Internet]. Vol. 26, NeuroImage. 2005. p. 441–53. Available from: 10.1016/j.neuroimage.2005.01.058

26. Hanley MA, Jensen MP, Ehde DM, Robinson LR, Cardenas DD, Turner JA, et al. Clinically significant change in pain intensity ratings in persons with spinal cord injury or amputation. Clin J Pain. 2006 Jan;22(1):25–31.

27. García-Rudolph A, Wright MA, Yepes C, Murillo N, Conesa L, Soriano I, et al. Effectiveness and efficiency of telerehabilitation on functionality after spinal cord injury: A matched case-control study. PM R. 2024 Aug;16(8):815–25.

28. Touchett H, Apodaca C, Siddiqui S, Huang D, Helmer DA, Lindsay JA, et al. Current Approaches in Telehealth and Telerehabilitation for Spinal Cord Injury (TeleSCI). Curr Phys Med Rehabil Rep. 2022 Apr 26;10(2):77–88.

29. Coulter EH, McLean AN, Hasler JP, Allan DB, McFadyen A, Paul L. The effectiveness and satisfaction of web-based physiotherapy in people with spinal cord injury: a pilot randomised controlled trial. Spinal Cord. 2017 Apr;55(4):383–9.

30. World medical association declaration of Helsinki. JAMA. 2013 Nov 27;310(20):2191.

31. Bouhassira D, Attal N, Alchaar H, Boureau F, Brochet B, Bruxelle J, et al. Comparison of pain syndromes associated with nervous or somatic lesions and development of a new neuropathic pain diagnostic questionnaire (DN4). Pain. 2005 Mar;114(1-2):29–36.

32. American College of Sports Medicine. ACSM’s Guidelines for Exercise Testing and Prescription. Wolters Kluwer; 2021. 513 p.

33. Qi L, Ferguson-Pell M, Salimi Z, Haennel R, Ramadi A. Wheelchair users’ perceived exertion during typical mobility activities. Spinal Cord. 2015 Sep;53(9):687–91.

34. Williams N. The Borg Rating of Perceived Exertion (RPE) scale. Occup Med (Lond). 2017 Jul;67(5):404–5.

35. de Oliveira BIR, Howie EK, Dunlop SA, Galea MP, McManus A, Allison GT. SCIPA Com: outcomes from the spinal cord injury and physical activity in the community intervention. Spinal Cord. 2016 Oct;54(10):855–60.

36. Rupp R, Schuld C, Biering-Sørensen F, Walden K, Rodriguez G, Kirshblum S, et al. A taxonomy for consistent handling of conditions not related to the spinal cord injury (SCI) in the International Standards for Neurological Classification of SCI (ISNCSCI). Spinal Cord [Internet]. 2021 Jun 9; Available from: 10.1038/s41393-021-00646-0

37. Ginis KAM, Martin Ginis KA, van der Scheer JW, Latimer-Cheung AE, Barrow A, Bourne C, et al. Evidence-based scientific exercise guidelines for adults with spinal cord injury: an update and a new guideline [Internet]. Vol. 56, Spinal Cord. 2018. p. 308–21. Available from: 10.1038/s41393-017-0017-3

38. Farrar JT, Young JP, LaMoreaux L, Werth JL, Poole MR. Clinical importance of changes in chronic pain intensity measured on an 11-point numerical pain rating scale. Pain. 2001 Nov;94(2):149–58.

39. Westaway MD, Stratford PW, Binkley JM. The patient-specific functional scale: validation of its use in persons with neck dysfunction. J Orthop Sports Phys Ther. 1998 May;27(5):331–8.

40. Keeney T, Slavin M, Kisala P, Ni P, Heinemann AW, Charlifue S, et al. Sensitivity of the SCI-FI/AT in Individuals With Traumatic Spinal Cord Injury. Arch Phys Med Rehabil. 2018 Sep;99(9):1783–8.

41. Slavin MD, Ni P, Tulsky DS, Kisala PA, Heinemann AW, Charlifue S, et al. Spinal Cord Injury–Functional Index/Assistive Technology Short Forms. Arch Phys Med Rehabil. 2016 Oct 1;97(10):1745–52.e7.

42. Penn RD, Savoy SM, Corcos D, Latash M, Gottlieb G, Parke B, et al. Intrathecal baclofen for severe spinal spasticity. N Engl J Med. 1989 Jun 8;320(23):1517–21.

43. Buysse DJ, Reynolds CF 3rd, Monk TH, Berman SR, Kupfer DJ. The Pittsburgh Sleep Quality Index: a new instrument for psychiatric practice and research. Psychiatry Res. 1989 May;28(2):193–213.

44. Krassioukov A, Biering-Sørensen F, Donovan W, Kennelly M, Kirshblum S, Krogh K, et al. International standards to document remaining autonomic function after spinal cord injury. J Spinal Cord Med. 2012 Jul;35(4):201–10.

45. Spielberger CD, Gorsuch RL, Lushene R, Vagg PR, Jacobs GA. Manual for the State-Trait Anxiety Inventory. Palo Alto, CA: Consulting Psychologists Press; 1983.

46. Kalpakjian CZ, Toussaint LL, Albright KJ, Bombardier CH, Krause JK, Tate DG. Patient health Questionnaire-9 in spinal cord injury: an examination of factor structure as related to gender. J Spinal Cord Med. 2009;32(2):147–56.

47. Roelofs J, van Breukelen G, Sluiter J, Frings-Dresen MHW, Goossens M, Thibault P, et al. Norming of the Tampa Scale for Kinesiophobia across pain diagnoses and various countries. Pain. 2011 May;152(5):1090–5.

48. Middleton JW, Tate RL, Geraghty TJ. Self-Efficacy and Spinal Cord Injury: Psychometric Properties of a New Scale. Rehabil Psychol. 2003 Nov;48(4):281–8.

49. Middleton JW, Tran Y, Lo C, Craig A. Reexamining the Validity and Dimensionality of the Moorong Self-Efficacy Scale: Improving Its Clinical Utility. Arch Phys Med Rehabil. 2016 Dec;97(12):2130–6.

50. Malouin F, Richards CL, Jackson PL, Lafleur MF, Durand A, Doyon J. The Kinesthetic and Visual Imagery Questionnaire (KVIQ) for assessing motor imagery in persons with physical disabilities: a reliability and construct validity study. J Neurol Phys Ther. 2007 Mar;31(1):20–9.

51. Alleva JM, Tylka TL, Kroon Van Diest AM. The Functionality Appreciation Scale (FAS): Development and psychometric evaluation in U.S. community women and men. Body Image. 2017 Dec;23:28–44.

52. Feng S, McDaniel S, Van de Winckel A. Finding functionality: Rasch analysis of the Functionality Appreciation Scale in community-dwelling adults in the US. Front Rehabil Sci. 2023 Oct 2;4:1222892.

53. Cramer H, Mehling WE, Saha FJ, Dobos G, Lauche R. Postural awareness and its relation to pain: validation of an innovative instrument measuring awareness of body posture in patients with chronic pain. BMC Musculoskelet Disord. 2018 Apr 6;19(1):109.

54. Dragesund T, Strand LI, Grotle M. The Revised Body Awareness Rating Questionnaire: Development Into a Unidimensional Scale Using Rasch Analysis. Phys Ther. 2018 Feb 1;98(2):122–32.

55. Carpentier S, Deng W, Blackwood J, Van de Winckel A. Rasch validation of the revised body awareness rating questionnaire (BARQ-R) in adults with and without musculoskeletal pain. BMC Musculoskelet Disord. 2024 Oct 9;25(1):799.

56. Mehling WE, Acree M, Stewart A, Silas J, Jones A. The Multidimensional Assessment of Interoceptive Awareness, Version 2 (MAIA-2). PLoS One. 2018 Dec 4;13(12):e0208034.

57. Tulsky DS, Kisala PA, Victorson D, Tate DG, Heinemann AW, Charlifue S, et al. Overview of the Spinal Cord Injury--Quality of Life (SCI-QOL) measurement system. J Spinal Cord Med. 2015 May;38(3):257–69.

58. Hall K, Dijkers M, Whiteneck G, Brooks CA, Krause JS. The Craig Handicap Assessment and Reporting Technique (CHART): Metric Properties and Scoring. Top Spinal Cord Inj Rehabil. 1998 Jul 1;4(1):16–30.

59. Van de Winckel A, Zhang L, Hendrickson T, Lim KO, Mueller BA, Philippus A, et al. Identifying body awareness-related brain network changes after Spring Forest QigongTM practice or P.Volve low-intensity exercise in adults with chronic low back pain: a feasibility Phase I Randomized Clinical Trial. medRxiv [Internet]. 2023 Mar 1; Available from: http://medrxiv.org/lookup/doi/10.1101/2023.02.11.23285808

60. Sevinc G, Hölzel BK, Hashmi J, Greenberg J, McCallister A, Treadway M, et al. Common and Dissociable Neural Activity After Mindfulness-Based Stress Reduction and Relaxation Response Programs. Psychosom Med. 2018 Jun;80(5):439–51.

61. Zeidan F, Vago DR. Mindfulness meditation-based pain relief: a mechanistic account. Ann N Y Acad Sci. 2016 Jun;1373(1):114–27.

62. Van de Winckel A, Carpentier S, Deng W, Zhang L, Battaglino R, Morse L. Using remotely delivered Spring Forest Qigong^TM^ to reduce neuropathic pain in adults with spinal cord injury: protocol of a quasi-experimental feasibility clinical trial. Pilot Feasibility Stud. 2023 Aug 22;9(1):145.

63. Burianová H, Marstaller L, Sowman P, Tesan G, Rich AN, Williams M, et al. Multimodal functional imaging of motor imagery using a novel paradigm. Neuroimage. 2013 May 1;71:50–8.

64. Batula AM, Mark JA, Kim YE, Ayaz H. Comparison of Brain Activation during Motor Imagery and Motor Movement Using fNIRS. Comput Intell Neurosci. 2017 May 4;2017:5491296.

65. Guillot A, Collet C, Nguyen VA, Malouin F, Richards C, Doyon J. Brain activity during visual versus kinesthetic imagery: an fMRI study. Hum Brain Mapp. 2009 Jul;30(7):2157–72.

66. Kilintari M, Narayana S, Babajani-Feremi A, Rezaie R, Papanicolaou AC. Brain activation profiles during kinesthetic and visual imagery: An fMRI study [Internet]. Vol. 1646, Brain Research. 2016. p. 249–61. Available from: 10.1016/j.brainres.2016.06.009

67. Lachenbruch PA, Cohen J. Statistical Power Analysis for the Behavioral Sciences (2nd ed.). J Am Stat Assoc. 1989 Dec;84(408):1096.

68. Whitfield-Gabrieli S, Nieto-Castanon A. *Conn*: A functional connectivity toolbox for correlated and anticorrelated brain networks. Brain Connect. 2012 Jun;2(3):125–41.

69. Asan AS, McIntosh JR, Carmel JB. Targeting sensory and motor integration for recovery of movement after CNS injury. Front Neurosci. 2021;15:791824.

70. Todd KR, Van Der Scheer JW, Walsh JJ, Jackson GS, Dix GU, Little JP, et al. The impact of sub-maximal exercise on neuropathic pain, inflammation, and affect among adults with spinal cord injury: A pilot study. Front Rehabil Sci [Internet]. 2021 Oct 26;2(700780). Available from: 10.3389/fresc.2021.700780

71. Boldt I, Eriks-Hoogland I, Brinkhof MWG, de Bie R, Joggi D, von Elm E. Non-pharmacological interventions for chronic pain in people with spinal cord injury. Cochrane Libr [Internet]. 2014 Nov 28;2014(11). Available from: 10.1002/14651858.cd009177.pub2

72. Rosner J, de Andrade DC, Davis KD, Gustin SM, Kramer JLK, Seal RP, et al. Central neuropathic pain. Nat Rev Dis Primers [Internet]. 2023 Dec 21;9(1). Available from: 10.1038/s41572-023-00484-9

